# Behavioral and Magnetoencephalographic Correlates of Fear Generalization Are Associated with Responses to Later Virtual Reality Exposure Therapy in Spider Phobia

**DOI:** 10.1101/2021.03.23.21253886

**Authors:** Kati Roesmann, Elisabeth Johanna Leehr, Joscha Böhnlein, Christian Steinberg, Fabian Seeger, Hanna Schwarzmeier, Bettina Gathmann, Niklas Siminski, Martin J. Herrmann, Udo Dannlowski, Ulrike Lueken, Tim Klucken, Kevin Hilbert, Thomas Straube, Markus Junghöfer

## Abstract

As overgeneralization of fear is a pathogenic marker of anxiety disorders, we investigated whether pre-treatment levels of fear generalization in spider-phobic patients are associated with their response to exposure-based treatment, in order to identify pre-treatment correlates of treatment success. Ninety patients with spider phobia completed pre-treatment clinical and magnetoencephalography (MEG) assessments, one session of virtual reality exposure therapy, and a post-treatment clinical assessment. Based on the primary outcome (30% symptom reduction in self-reported symptoms from pre-to post-treatment) they were categorized as responders or non-responders. In a pre-treatment MEG fear generalization paradigm involving fear conditioning with two unconditioned stimuli (UCS), we obtained fear ratings, UCS-expectancy ratings, and event-related fields to conditioned stimuli (CS+, CS-) and 7 different generalization stimuli (GS) on a perceptual continuum from CS+ to CS-. Prior to treatment, non-responders showed behavioral overgeneralization indicated by more linear generalization gradients in fear ratings. Analyses of MEG source estimations revealed that non-responders showed a decline of their (inhibitory) frontal activations to safety-signaling CS- and GS compared to CS+ over time, while responders maintained these activations at early (<300ms) and late processing stages. Results provide initial evidence that pre-treatment differences of behavioral and neural markers of fear generalization are associated with later responses to behavioral exposure. Findings demonstrate the relevance of inhibitory learning functions and their spatio-temporal neural reflections in this interplay. Findings stimulate research on mechanism-based augmentation strategies for behavioral therapies.

## 1. Introduction

The first-line treatment for anxiety disorders is exposure-based cognitive-behavioral therapy [1]. Yet, not all patients benefit equally. Non-response rates of up to 50% [2] have stimulated research on factors that moderate treatment outcomes [3]. Despite considerable heterogeneity of study approaches and findings, most identified factors are in line with the inhibitory learning framework of exposure therapy (ET) [4]. This model assumes that ET does not erase the fear-associated relations between phobic stimuli (conditioned stimuli, CS; e.g., a spider) and expected aversive outcomes (unconditioned stimuli, UCS; e.g., a bite) – i.e., the CS/UCS traces. Instead, it posits that ET produces new co-existing inhibitory CS/noUCS traces that connect the CS with safety information [5, 6]. Thus, reductions of fear are expected if patients learn to successfully discriminate between danger and safety signals and to inhibit fear responses in the presence of safety cues [7].

Accordingly, one pre-treatment factor that might account for patients’ variability in responses, is the variance in inhibitory learning functions which can be investigated in classical conditioning experiments. Aberrant processes during fear acquisition and extinction [8, 9], as well as fear generalization [10–13] are considered promising pathogenic markers of anxiety disorders. Fear generalization is defined as the transfer of conditioned fear responses to so-called generalization stimuli (GS) that have never been paired with the UCS but that share similarity with the threat-signaling CS+, such as orientation [14], size [10] or shape [15]. The degree of fear generalization can be described by a generalization gradient along the relevant dimension from the safety-signaling CS-via GS to the threat-signaling CS+.

Thereby, more fear generalization, or overgeneralization of fear, is indicated by linear (vs. quadratic) and shallower gradients. Compared with healthy controls, this phenomenon has been observed for patients with various anxiety disorders [10, 11, 16], but see [17]. Recent fear conditioning studies have strengthened the hypothesis, that inhibitory learning mechanisms contribute to the degree of fear generalization: For example, brain activity in frontal networks that support fear inhibition [18] *decreases* as the GS approximates the threat-signaling CS+ [15, 19–21]. Such negative (“inhibitory”) gradients, which peak at or near the CS-, seem to be modulated by pathological anxiety, with shallower negative generalization gradients in the ventromedial prefrontal cortex (vmPFC) in generalized anxiety disorder [17]. Such failure to recruit relevant brain networks supporting fear inhibition in the presence of safety cues likely underpins anxiety-related overgeneralization (for review see [12]).

Negative generalization gradients have also been reported for the dorsolateral prefrontal cortex (dlPFC), a region which also supports inhibitory processes during behavioral exposure [22]. Employing highly temporally resolving magnetoencephalography (MEG) which allows to estimate underlying neural sources of event-related fields (ERF) with an acceptable spatial resolution, we recently showed stronger “inhibitory” dlPFC activity evoked by GS that resemble the safety signaling CS-[23]. These effects occured at rather late time intervals (>300ms), reflecting strategic emotion-regulation [24], but also already within the first 100ms of stimulus processing.

Several studies have linked behavioral and neural pre-treatment correlates of fear acquisition and extinction with treatment outcomes in ET [25–27]. However, such a relationship has not been examined for fear generalization. Therefore, we here investigated whether pre-treatment mechanisms of fear generalization are associated with later responses to ET. We hypothesized that later non-responders versus responders would show pre-treatment overgeneralization in their behavioral and neural (MEG) fear responses. On a neural level, non-responders’ overgeneralization should particularly be reflected in shallower negative frontal gradients. To delineate temporal characteristics of associations between generalization gradients and treatment-response to ET, we separately tested early (<300ms) and late stages (>300ms) of processing within anterior neural networks. Predictions were tested in a sample of spider phobic patients, who underwent a well-controlled virtual reality exposure therapy (VRET) [28].

## 2. Methods and Materials

### 2.1 Participants

Ninety patients with spider phobia according to the structured clinical interview for DSM-IV [29] completed pre-treatment clinical and magnetoencephalography (MEG) assessments, one session of virtual reality exposure therapy, and a post-treatment clinical assessment (Figure 1). Patients were aged between 18 and 65 years, of Caucasian decent, right-handed and fluent in German (Table 1). This study was approved by the Ethics Committee of the Medical Faculty of the University of Münster. Patients consented to participate in all assessments and were compensated with 180€. For more details, see SM1.1.

**Table 1:** Demographic and clinical characteristics of treatment responders and non-responders before and after VRET for the MEG sample (N = 70).

| Characteristic | Responders <sup>#</sup> | | Non-Responders <sup>##</sup> | | Test<br>t [$\chi^2$] | df | p |
| --- | --- | --- | --- | --- | --- | --- | --- |
|  | M | SD | M | SD |  |  |  |
| **Primary Response Criterion** |  |  |  |  |  |  |  |
| (Reduction in SPQ [%]) | >= 30 |  | < 30% |  |  |  |  |
| Sample size [N, %] | 43/89 | 48.31 | 46 | 51.69 |  |  |  |
| Reduction in SPQ [%, SD] | 42.06 | 8.41 | 20.66 | 6.78 | 13.258 | 87 | <.001*** |
| **Demographic Characteristics** |  |  |  |  |  |  |  |
| Female gender [N, %] | 35 | 81.4 | 36 | 78.3 | $\chi^2 = 1.35$ | 1 | .713 |
| Age (years) | 26.07 | 7.81 | 28.85 | 9.24 | 1.527 | 87 | .130 |
| Years of education | 14.91 | 2.57 | 14.91 | 2.89 | 0.007 | 86 | .994 |
| **Clinical Characteristics** |  |  |  |  |  |  |  |
| Age of onset (years) | 5.76 | 4.45 | 6.73 | 5.15 | 0.590 | 86 | .557 |
| Comorbid major depression [N, %] | 3 | 7.0 | 1 | 2.2 |  |  |  |
| Comorbid subordinate animal phobia [N, %] | 1 | 2.3 | 1 | 2.2 |  |  |  |
| SPQ pre | 22.95 | 2.03 | 22.28 | 1.93 | 1.697 | 87 | .114 |
| SPQ post | 13.26 | 2.00 | 17.67 | 2.12 | 10.09 | 87 | <.001*** |
| BAT (final distance in cm) pre | 173.71 | 76.91 | 171.97 | 75.11 | 0.108 | 87 | .914 |
| BAT (final distance in cm) post | 78.38 | 67.32 | 98.40 | 70.62 | 1.367 | 87 | .175 |
| Reduction in BAT [%, SD] | 56.04 | 32.35 | 41.65 | 31.08 | 2.140 | 87 | .035* |
| CGI pre [N, %] | | | | | $\chi^2 = 2.717$ | 3 | .437 |
| Mildly ill | 4 | 9.3 | 6 | 13.0 |  |  |  |

| Characteristic | Responders <sup>#</sup> |  | Non-Responders <sup>##</sup> |  | Test |  |  |
| --- | --- | --- | --- | --- | --- | --- | --- |
| | M | SD | M | SD | t [$\chi^2$] | df | p |
| Moderately ill | 24 | 55.8 | 18 | 39.1 |  |  |  |
| Markedly ill | 15 | 34.9 | 19 | 41.3 |  |  |  |
| Severely ill | 0 | 0.0 | 1 | 2.2 |  |  |  |
| CGI post [N, %] | | | | | $\chi^2 = 6.615$ | 4 | .158 |
| Not ill | 2 | 4.7 | 1 | 2.2 |  |  |  |
| Marginally ill | 5 | 11.6 | 2 | 4.3 |  |  |  |
| Mildly ill | 24 | 55.8 | 20 | 43.5 |  |  |  |
| Moderately ill | 9 | 20.9 | 21 | 45.7 |  |  |  |
| Markedly ill | 2 | 4.7 | 2 | 4.3 |  |  |  |
| Severely ill | 0 | 0.0 | 0 | 0.0 |  |  |  |
| STAI trait pre | 35.58 | 9.69 | 36.022 | 8.21 | 0.231 | 86 | .818 |
| BDI-II total pre | 3.70 | 4.74 | 3.58 | 3.95 | 0.129 | 86 | .898 |
| BDI-II total post | 4.05 | 5.75 | 3.07 | 3.34 | 0.990 | 86 | .325 |
| UI-18: total pre | 41.16 | 13.51 | 39.11 | 12.52 | 0.739 | 86 | .462 |
| UI-18: total post | 42.05 | 13.42 | 41.63 | 12.97 | 0.148 | 86 | .883 |

*Note.* Means, standard deviations, t-values (M, SD, t; except where noted), degrees of freedom (df) and significance level (p, two-sided) for each characteristic; SPQ, Spider Phobia Questionnaire; BAT, behavioral avoidance test; CGI, Clinical Global Impression; STAI-Trait, trait-version of the State-Trait Anxiety Inventory; BDI-II, Beck Depression Inventory-II; UI-18, Intolerance of Uncertainty Scale – 18. # Missing values for responders: Age of onset (1), CGI rating post (1), BDI post (1), Ui-18: total post (1); Imputed data for non-responders (0) ## Missing values for non-responders: Years of education (1), CGI rating pre (2), STAI pre (1), BDI pre (1), UI-18: total pre (1); Imputed data for non-responders: BAT pre (1), BAT post (1). Characteristics of the MEG sample (N=70) as well as references of the employed psychometric questionnaires are presented in Table S1.

**Figure 1:**
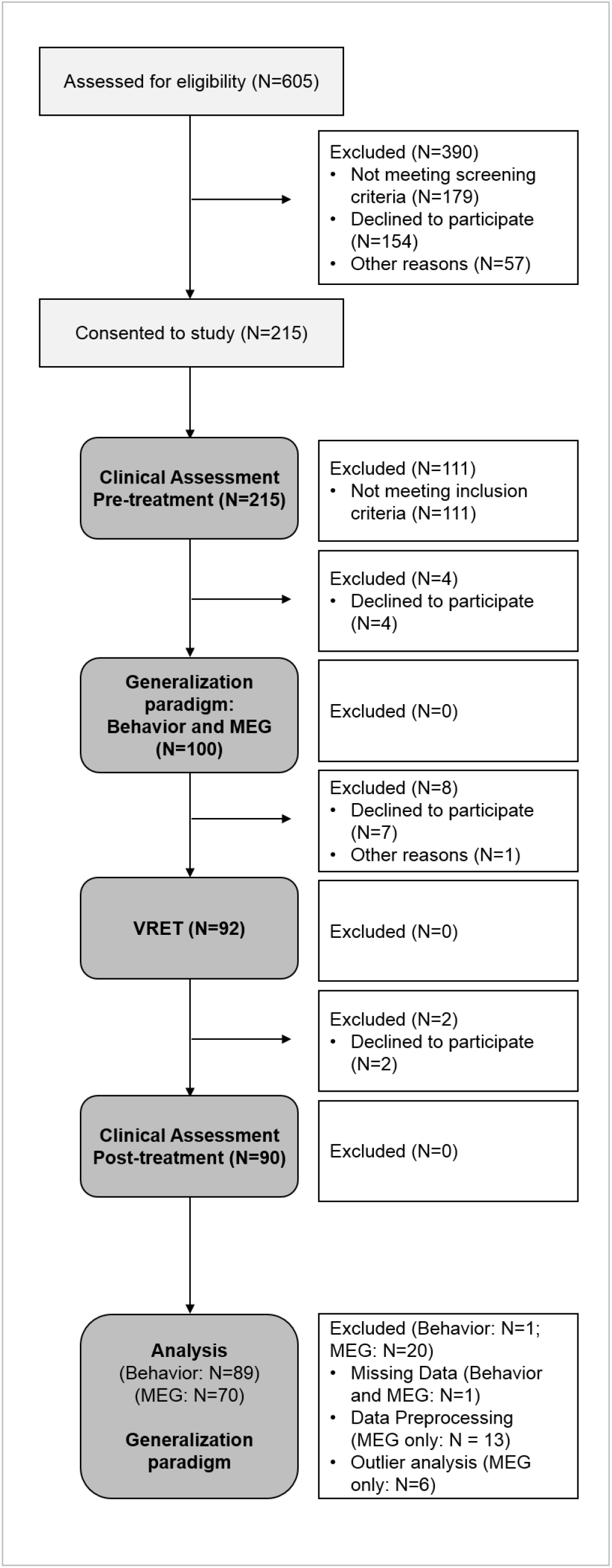
Flow-chart visualizing the recruitment pathway of the clinical sample described here. 100 eligible patients completed behavioral and magnetoencephalographic assessments on fear generalization. Out of these, 92 patients completed the virtual reality exposure therapy (VRET) and 90 completed the clinical post-treatment assessment. One of these patients was excluded from the behavioral and the MEG-analysis, because one part of the generalization paradigm was not completed. Another 19 patients were excluded from the MEG-analyses due to insufficient data quality in any MEG acquisition (preprocessing criterion: N=13, outlier analysis: N=6). Note that this study was embedded in a joint prospective longitudinal project of the Transregional Collaborative Research Center “Fear, Anxiety, Anxiety Disorders” (CRC-TRR58 funded by the German Research Foundation) employing VRET as a first-line treatment for specific phobia and pre-defined response criteria (for details, see supplemental materials: SM1).

### 2.2 Material

#### 2.2.1 Clinical Assessment of Treatment-Response

We employed percentual reductions from the clinical pre-to post-treatment assessment in the Spider Phobia Questionnaire (SPQ [30], registered as primary outcome at Clinical Trials.gov (NCT03208400) to determine categorical treatment-response (TR-cat, responders vs. non-responders) and dimensional outcomes (TR-dim). Responders were characterized by SPQ-reductions of >30%, i.e. a clinically meaningful response (see [28]).

#### 2.2.2 Behavioral and MEG Assessment on Fear generalization

##### Conditioned Stimuli and Generalization Stimuli (CS and GS)

Four sets of sinusoidal grating stimuli, each consisting of 9 stimuli, served as CS and GS. The two most different stimuli in each set differed by 24° in their tilt angles and were used as CS+ and CS-(orientations: 11°/35°, 101°/125°, 56°/80°, and 146°/170°). Seven additional GS ranged in steps of 3° between each CS+ and CS-pair. The assignment of CS+/CS-orientations was balanced.

##### Unconditioned Stimuli (UCS)

A picture of a spider and a picture of a threatened female face [31] were selected as phobia-related and phobia-unrelated UCS. Two auditory stimuli (a female scream [32] and a white noise; 60 dB above individual hearing threshold) were presented simultaneously with the visual stimuli, in counterbalanced combinations across participants and blocks (see Figure 2a). To match physical characteristics the white noise was multiplied with the scream’s temporally inverted amplitude curve. Fear ratings were higher for phobia-related compared to phobia-unrelated UCS and did not differ between responders and non-responders (see SM2.2.1).

**Figure 2:**
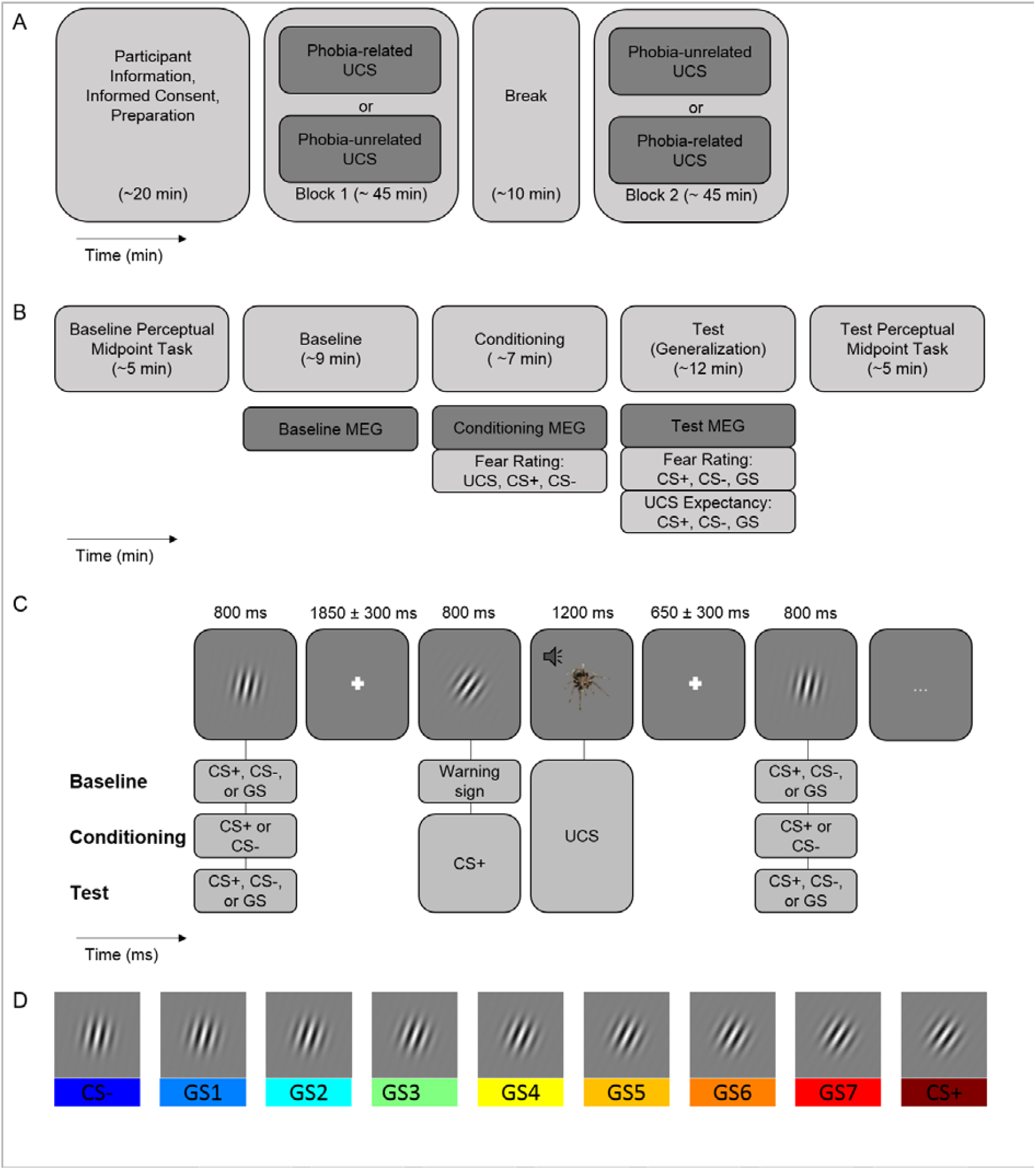
Experimental Procedure for the Behavioral and MEG Assessment on Fear generalization. A) Overview on the procedure, consisting of an informatory block and two experimental blocks that resembled each other except for randomization and the applied *UCS-type* (phobia-related vs phobia-unrelated). The order of blocks was counterbalanced across patients. B) Each block consisted of a Baseline Perceptual Midpoint (PM) Task, an MEG-Baseline phase, an MEG-Conditioning phase which terminated with a fear rating of the CS+, the CS- and the respective audiovisual USs, an MEG-Test-Phase, which terminated with fear- and UCS expectancy ratings of the CS+, the CS- and all GS, and – again – the PM Task. C) Sequence of stimulus presentation during the MEG baseline, conditioning and test phase in a phobia-related block. Stimuli were repeatedly presented. In the Conditioning and Test phase the CS+ predicted the US (here: phobia related) in 33% of the cases while a warning signal predicted all US in the baseline phase. Parallel to the MEG-signal pupil dilation was recorded. D) Example of stimuli used as CS and GS (here: set A, orientations between 11° and 35°).

### 2.3 Procedure

Patients first underwent a clinical pre-treatment assessment, which included primary and secondary outcome measures, demographic and psychometric questionaires, followed by the behavioral and MEG assessment on fear generalization (+13.03 days, SD=9.38), the VRET (+27.46 days, SD=10.35), and the clinical post-treatment assessment (+M=32.70, SD=12.02, see Figure 1, for details see [28] and SM1.2-1.2.2). During VRET, patients were exposed to virtual spiders in up to five different scenarios. The duration of behavioral exposure (t(87)=1.247, p=.216, M = 77.69 min, SD = 22.79) and the experienced immersion in the virtual environment measured by the Igroup Presence Questionnaire (IPQ, [33]) did not differ between responders and non-responders (p’s in all subscales >.16, General Presence [0;6]: M=4.91, SD=0.95], Spatial Presence [0;30]: M=20.74, SD=12.63; Involvement [0;24]: M=13.60, SD=11.84, Experienced Realism [0;24]: M=12.31, SD=3.71).

#### 2.3.1 Behavioral and MEG Assessment on Fear generalization

The experiment consisted of two similar blocks that differed only in randomization and in the presented *UCS-type* (phobia-related and phobia-unrelated as with-in subject factor). Each block consisted of a Baseline-MEG phase, a Conditioning-MEG phase that ended with a fear rating of CS+, CS- and the respective UCS, and a Test-MEG phase that ended with fear- and UCS expectancy ratings of all CS+, CS- and GSs. In a subsample, pupil dilation to CS and GS was recorded in parallel with ERFs. In a Baseline and Test Perceptual Midpoint (PM) task before and after MEG assessments (see Figure 2A and B) participants indicated whether GSs were perceptually more similar to the CS+ or the CS-. Results revealed that discrimination abilities and perceived perceptual midpoints of GSs were not modulated by phase (baseline vs. test), by treatment-response or by interactions of these factors (see SM1.2.3. and SM2.2.3)

In each phase of MEG- (and pupil) recordings (see Figure 2, paradigm based on [20]), CS+, CS-, and the 7 GSs (in baseline and test phases only) were presented consecutively in the center of the screen for 800ms with a random inter-stimulus-interval of 1550-2150ms. Participants kept their attention on the presented stimuli while behavioral reactions were not required.

In the Baseline- and Test-phases, all CS+, CS- and GS were presented 21 times each in pseudorandomized order. At baseline, we recorded responses to all 9 *stimulus-types* (within-subject factor) *before* they were contingently associated with the UCS. Additionally, the respective UCS (phobia-related or unrelated) was presented seven times and was – as instructed – always preceded by a warning sign (square or triangle, contingency rate: 100%) to achieve constant arousal levels across baseline- and test-phases (21). In the conditioning-phase, CS+ and CS-stimuli were presented 60 times each in pseudorandomized order, whereby the UCS (known from the baseline phase) replaced 33% of CS+. Patients were instructed that one grating with a specific orientation was UCS predictive. Patients then rated the level of fear elicited by the CSs and UCSs by button press on a ten-step numeric rating scale ranging from “no fear” to “extreme fear” (i.e. fear ratings; instruction: “How much fear does this grating elicit?”). In the subsequent Test-phase, responses to all *stimulus-types after* conditioning were assessed. CS+ continued to predict by the UCS (contingency rate: 33%, 7 UCS presentations) to prevent extinction [34]. Again patients were fully instructed beforehand. Finally, behavioral fear- and UCS-expectancy of CS+, CS-, and GS (three repetitions per stimulus; pseudorandomized order) ratings were obtained. For UCS-expectancy ratings, patients estimated the probability the visually presented *stimulus-type* predicted the UCS on a ten-step numeric rating scale (“very unlikely” to “very likely”; instruction: “How likely was it, that this grating was followed by a spider/face?”).

#### 2.3.2 Recording and Pre-processing of MEG Data

ERFs were recorded and pre-processed as described in our previous MEG study on fear generalization [23]. The spatiotemporal signal to different stimulus-types was computed separately for each participant, block and phase. To increase the number of trials per category and thus the signal-to-noise ratio, we merged ERFs in reaction to neighboring gratings (i.e., moving average) as previously applied [10]. Cortical sources underlying the averaged ERFs were estimated using the L2-Minimum-Norm-Estimates (L2-MNE) method [23].

To avoid statistical artifacts due to potential outliers, patients were excluded if the mean of the standard deviation across time between experimental conditions (N=6) or the mean number of residual trials across experimental conditions (N=0) differed from the sample median by more than four standard deviations. For visualization purposes, L2-MNE topographies were projected on standard 3D brain models.

### 2.4 Analysis

To test the hypothesis that pre-treatment generalization gradients are related to the later *treatment-response*, we specifically explored interactions of *treatment-response* with linear and quadratic contrasts in the factor *stimulus-type* and *stimulus-type* by *UCS-type* interactions using mixed-measures ANOVAs (for TR-cat) and ANCOVAS (for TR-dim). Analyses were run separately for fear and UCS-expectancy ratings (obtained in the test-phase), and estimated neural activity (L2MNE). Additionally, fear ratings of CS and UCS (obtained post-conditioning) were analysed using ANOVAS. Clinical and behavioral data were analyzed using the programs R (2015) and SPSS 24 (IBM Corp., Armonk, NY) with a significance level of α=.05.

To focus on gradients specifically induced by the conditioning procedure and to extract purely perceptual effects elicited by the different tilt angles of the stimuli, all statistical MEG analyses were based on difference topographies (Test minus Baseline). First, we aimed to identify spatiotemporal clusters reflecting linear generalization effects observed in the fear and UCS-expectancy ratings of responders and non-responders. Thus, linear contrasts with the factor *stimulus-type* across groups were calculated for each time point and dipole. Identified clusters that additionally revealed treatment-outcome-dependent (TR-cat and TR-dim) modulations of gradients (Bonferroni-corrected for the number of clusters) are presented in the main text. Second, to directly identify clusters displaying differential generalization of responders and non-responders, difference topographies were investigated in a *stimulus-type* by *treatment-response* interaction analysis, in which orthogonal linear contrasts [17] were calculated for each time point and dipole^1^.

To correct for multiple comparisons within the predefined time intervals (IOI: 0-300ms, 300-600ms) and anterior region of interest (ROI), we adopted a non-parametrical statistical testing procedure [35] as used previously [23] to determine significant spatiotemporal clusters. Details on recording, preprocessing and analyses of MEG and pupil responses as well as supplementary analyses on dimenstional outcomes and individual response prediction are presented in SM 1.2.4-7.

## 3. Results

### 3.1 Clinical effect of VRET

SPQ-reductions from pre-to post-treatment assessment (t(88)=20.59, p<.001, d=2.18, M_Pre_ = 22.61, SD_Pre_ = 2.00; M_Post_ = 15.54, SD_Post =_ 3.02) supported the expectation of a highly effective VRET (for clininical effects, see also [36] and SM2.1). According to our primary outcome criterion (30% SPQ-reduction [28]), 48.31% of all patients responded to VRET (SPQ-reductions of 42.06% in responders and 20.66% in non-responders). SPQ-responders were also characterized by stronger percentual pre-to-post reductions of avoidance in the BAT than non-responders (t(87)=2.140, p=.035). Importantly, responders and non-responders did not differ in primary (SPQ) and secondary (BAT) outcome measures before therapy (for details see Table 1).

### 3.2 Fear Ratings and UCS-Expectancy Ratings

Post-conditioning, but before the test phase, we revealed different fear ratings of CS+ and CS- and a modulation of this differentiation by *UCS-type*, while CS+, CS- and UCS fear ratings did not differ between responders and non-responders (see SM2.2.1).

After the test phase, we observed a significant linear (F(1,87)=146.704, p<.001, η^2^=.628) and a quadratic (F(1,87)=3.984, p=.049, η2=.044) main effect for *stimulus-type*, with flattened gradients for GS showing increasing similarity with the CS-(see Figure 3, top). As predicted, we found a significant interaction of *stimulus-type* by *treatment-response*, which was evident for the quadratic (F(1,87)=5.046, p=.027, η^2^=.055) but not for the linear gradient (F(1,87)=0.014, p=.906, η^2^=.000) and independent of *UCS-type* (F(1,87)=0.836, p=.363, η^2^=.010). Post-hoc analyses for each group revealed quadratic gradients in responders (F(1,42)=7.317, p= .010, η^2^=.148) but not in non-responders (F(1,45)=0.039, p= .844, η^2^=.001). UCS-expectancy ratings also showed significant linear (F(1,87)=288.441, p<.001, η^2^=.768) and quadratic (F(1,87)=32.498, p<.001, η^2^=.272) main effects of *stimulus-type*. However, UCS-expectancy ratings did not differ between responders and non-responders (linear: F(1,87)=0.652, p=.421, η^2^=.007; quadratic: (F(1,87)=0.199, p=.657, η^2^=.002, see Figure 3, bottom). The three-way interaction *stimulus-type* by *treatment-response* by *UCS-type* was not significant (linear: F(1,87)=0.121, p=.729, η^2^=.001; quadratic: F(1,87)=0.561, p=.456, η^2^=.006). Associations between treatment-response and quadratic gradients in the factor *stimulus-type* of fear ratings could be confirmed in the MEG-sample for both TR-cat and TR-dim (see SM2.2.2, Table S2). Pupil responses revealed the expected linear positive gradients which were, however, not modulated by *treatment-response* (see SM2.3).

**Figure 3:**
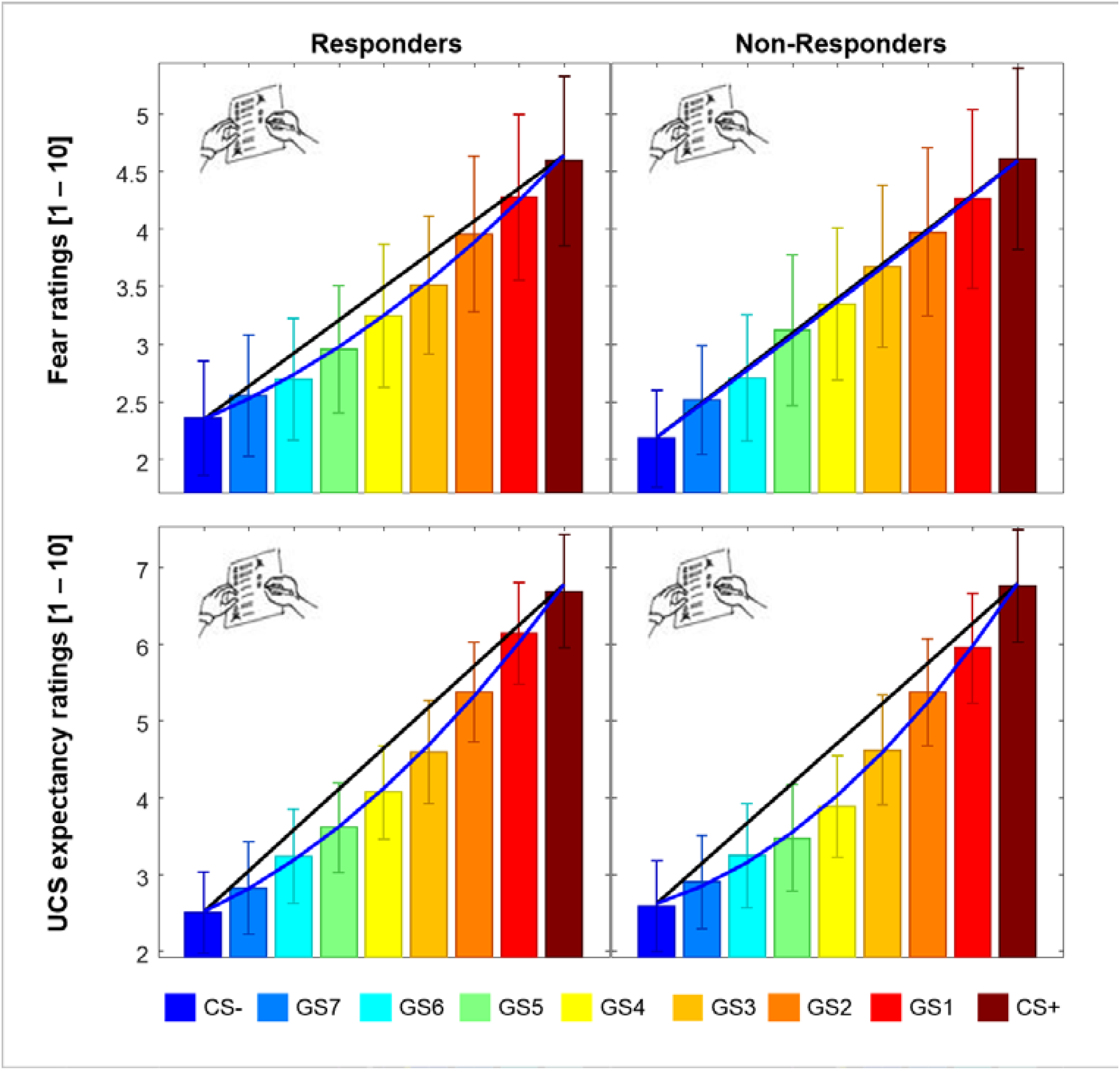
Results of fear and UCS-expectancy ratings. Linear (black line) and quadratic (blue line) behavioral generalization effects in fear rating and in UCS-expectancy ratings (N=89) are indicated. Fear ratings revealed overgeneralization in non-responders as indexed by a missing quadratic gradient in this group, i.e. a significant interaction in quadratic gradients. UCS-expectancy ratings revealed equivalent quadratic and linear grandients in both groups. Note that fear ratings and UCS-expectancy ratings revealed qualitatively similar results in the MEG sample of N=70 patients (see Table S2).

### 3.3 MEG data

#### 3.3.1 Negative “inhibitory” anterior linear contasts

Overall four anterior clusters that were characterized by a negative gradient peaking at the safety-signaling CS-^2^ yielded significance (see SM2.4.1., Figure S2). One left dlPFC cluster (Figure 4A, 110-157ms after stimulus onset, p-cluster=0.003) additionally yielded a significant interaction of *stimulus-type* by *treatment-response* (F(1,68)=9.351, p=.003, p_corrected_ =.012, η^2^=.121) that was qualified by a distinct negative linear gradient in responders (F(1,35)=26.830, p<.001, η^2^=.434) but not in non-responders (F(1,33)=1.900, p=.177, η^2^=.054). Again, treatment-response-dependent gradients were not modulated by *UCS-type* (F(1,68)=1.205, p=.276, η^2^ =.017). Convergent to the categorical analyis, linear gradients in this cluster were also associated with TR-dim (F(1,68) = 6.616, p =.012, p_corrected_=.048, η^2^ = 0.089, Table S3). TR-dim was additionally associated with the quadratic gradient in a cluster in right ventrolateral prefrontal regions that extended to the right TPJ (210-300ms, F(1,68) = 7.499, p = .008, p_corrected_= .032, η^2^=.099). Specifically, larger percentual reductions of SPQ-scores were associated with stronger negative quadratic gradients. This pattern resulted from overall stronger brain responses to the CS- and GS compared to the CS+ (see Figure S4). Negative linear contrasts at late latencies or positive linear contrasts were non-signficant in anterior regions.

**Figure 4:**
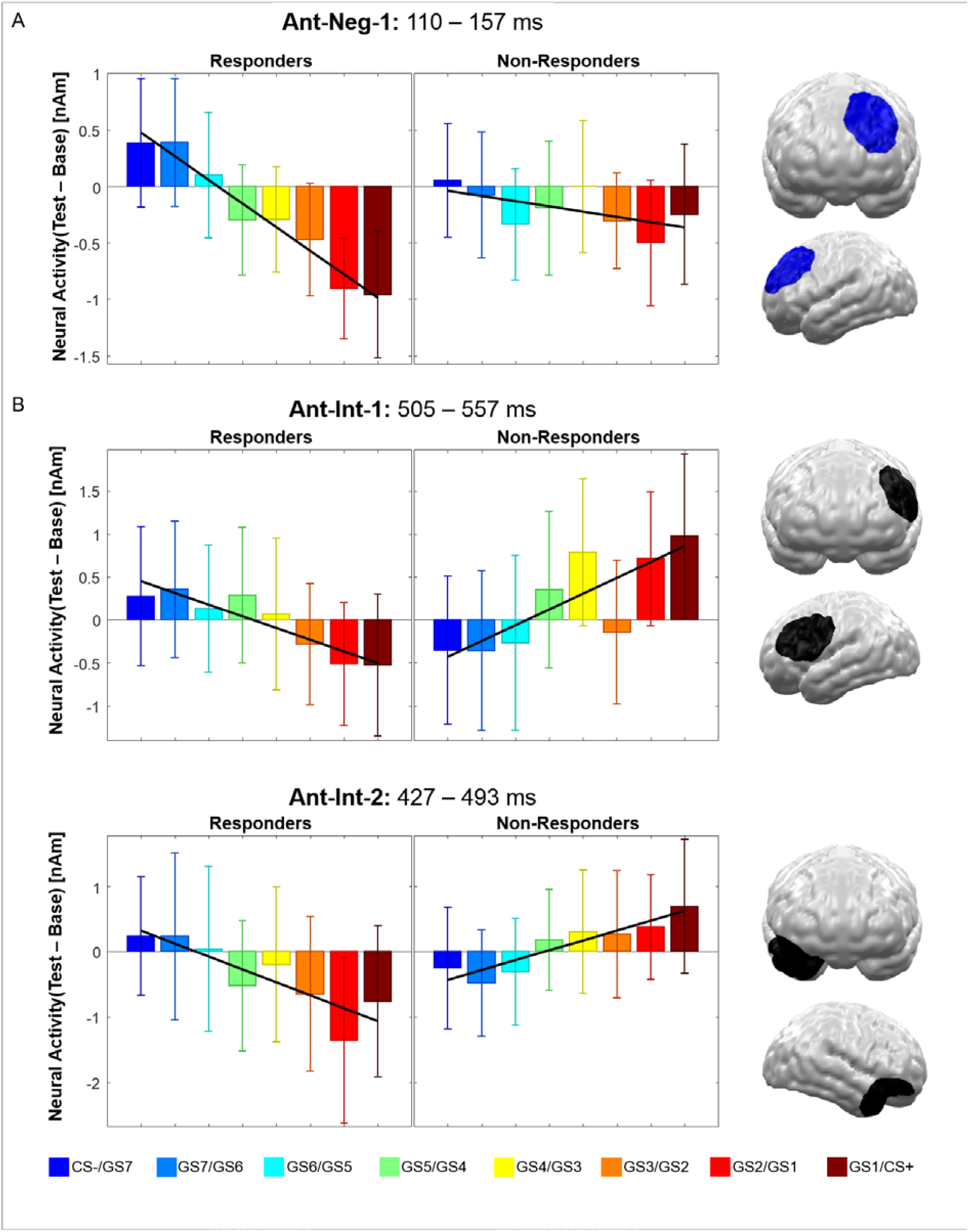
Significant spatiotemporal clusters showing differential generalization gradients for responders and non-responders in the anterior region of interest (Ant). (A) Cluster Ant-Neg 1 revealed a significant “inhibitory” gradient in the permutation test (Neg, indicated in blue) at early latencies. Importantly, while responders showed stronger brain activations to generalization stimuli resembling the safety-signaling CS-, non-responders showed no differentiation. (B) Significant spatiotemproal clusters revealing interaction effects (Int) in linear gradients, i.e. orthogonal contrasts (indicated in black), were revealed for the late time interval only. Both clusters revealed negative linear gradients for responders, while non-responders revealed the opposite pattern. The observed clusters spatially overlap with regions showing main effects in the early time interval (Figure S2). Bar graphs show the regional neural activity (Test minus Base in nAM) in the displayed clusters. Error bars denote 95% confidence intervals.

#### 3.3.2 Orthogonal linear contrasts

Orthogonal linear contrasts (a complementary analysis directly testing for associations of linear gradients and *treatment-response* (TR-cat) without necessity of negative gradients across groups) revealed two additional clusters at rather late latencies (see Figure 4B).

In a left dorso- and ventrolateral prefrontal cluster (505–557ms, p-cluster=.025), responders showed a marginal negative gradient (F(1,35)=3.907, p=.056, *η*^*2*^ =.100) while non-responders revealed a significant positive gradient (F(1,33)=10.698, p =.003, *η*^*2*^=.245). Similar effects were observed in a cluster spanning areas of the right anterior temporal pole and ventral orbitofrontal cortex (427–493ms, p-cluster=.013) where responders again revealed negative (F(1,35)=11.937, p=.001, *η*^*2*^ =.254) but non-responders positive gradients (F(1,33)=7.993, p=.008, *η*^*2*^=.195). Again, effects within both clusters were not modulated by the *UCS-type* (Fs<1). Supplemental analyses based on the dimensional treatment outcome (TR-dim, SM2.4.3-4) further confirmed these results. First, ANCOVAS within the two orthogonal clusters revealed that linear gradients in both clusters were also significantly associated with TR-dim (Table S4). Second, cluster permutation analyses on associations between individual linear polynomial parameters (per dipole and timepoint) and TR-dim in the anterior ROI were performed. These regression analyes yielded one cluster in bilateral anterior temporal areas and large areas of orbital and vmPFC (Figure S5) that covered areas of cluster Ant-Int-2 and supported the link between stronger “inhibitory” frontal negative gradients and symptom reductions. This analysis yielded no clusters in left dlPFC/vlPFC.

### 3.4 Prediction of individual responses to treatment – an exploratory machine learning approach

With a supplemental machine learning analysis, we estimated the potential of individual generalization gradients as predictors for individual treatment-responses (TR-cat), rather than a group-level correlate thereof [37]. Our prediction model, which included linear and quadratic polynomial coefficients of fear and expectancy ratings, and of neural activity in clusters revealing linear gradients as features, revealed a mean balanced prediction accuracy of 58.6% (p = .076, SM2.5).

## 4. Discussion

We aimed to test the hypothesis that pre-treatment mechanisms of fear generalization in anxiety patients and related inhibitory neural mechanisms in PFC regions are associated with later treatment outcomes to behavioral exposure therapy. We found that spider-phobic patients who did not respond to later VRET showed an overgeneralization in their behavioral fear ratings as well as diminished inhibitory or even inverted gradients in frontal brain networks compared to treatment-responders. These findings are in line with current models of inhibitory learning as a core mechanism for the development, maintenance and treatment of anxiety disorders. They furthermore show that pre-treatment configurations in brain networks conferring fear generalization are related with treatment-response, thus potentially opening up new avenues for personalized treatment augmentation (e.g. degeneralization trainings).

Both groups showed the expected linear gradients in fear ratings, but responders only additionally followed a quadratic trend indicative of less generalization. This finding suggests that deficits of fear generalization in later non-responders compared to responders resemble aberrant fear generalization of anxiety-disordered patients compared to healthy controls [10, 11]. Importantly, responders and non-responders did not differ regarding their baseline fear levels or further potential confounds like evaluations of CS and UCS (SM2.2.1), perceptual aspects of fear conditioning assessed in the PM task (SM1.3.3), relevant psychometric variables or treatment characteristics like duration of exposure or experience of immersion. Further, effects were specific for fear ratings and not evident in UCS expectancies or pupil dilation^3^. The dissociations of fear- and expectancy ratings suggest that affective, rather than cognitive aspects [34] of fear generalization are linked with treatment success.

As predicted, negative linear gradients were observed in dorso- and ventrolateral prefrontal networks [23] and revealed relatively stronger reactions to safety-signaling CS- and GS (vs. CS+) in responders. This replication of early-onset negative gradients in dorsolateral prefrontal regions [23] further emphasizes that “inhibitory” frontal structures – amongst other functions – modulate early, presumably more implicit aspects of fear generalization. Importantly, the early left-hemispheric dlPFC negative gradients were driven by the later treatment-responders, while non-responders revealed no inhibition of safety cues. A convergent, even more pronounced interaction was found in left-lateralized dlPFC/vlPFC in a later time interval (505–557ms). Correspondingly, dimensional treatment outcomes were associated with linear polynomial parameters in all of these left dlPFC/vlPFC clusters, and with quadratic parameters in the right vlPFC. Overall, this pattern suggests that responders maintained inhibitory frontal activations to safety-signaling CS- and GS compared to CS+ across time, while non-responders failed to do so. In combination with the behavioral evidence for overgeneralization of fear in non-responders, this observation supports previous speculations that overgeneralization might be mediated by aberrant dlPFC recruitment [38]. Importantly, the dlPFC supports regulatory processes, which are needed to differentiate relevant stimuli and to overcome anxiety in the presence of safety [38]. In line with this, influential models of emotion regulation converge in the notion of inhibitory top-down influences of lateral prefrontal structures on neural systems involved in emotion generation (e.g. amygdala, [18]) and perception (see SM2.4.5). Linking negative frontal generalization gradients with treatment-response to VRET, our study provides initial support for the hypothesis that inhibitory learning during exposure therapy [4, 22] and during fear generalization are mediated by at least overlapping regulatory brain systems. The finding of fast, emotional correlates of inhibitory deficiencies in non-responders suggests that implicit affective processes might be a promising target for therapeutic augmentation strategies (e.g. degeneralization training), and might complement cognitive and context-based strategies suggested in inhibitory learning models [4].

Yet, several aspects require consideration: First, the categorical analysis of treatment response did not reveal the predicted generalization gradients and/or differences of gradients between responders and non-responders in the vmPFC, which has often been associated with fear-inhibition during safety learning [15, 19–21] and overgeneralization of fear in anxiety disorders [12, 17]. However, supplemental dimensional analyses in fact revealed an increasingly improved treatment outcome with increasingly negative “inhibitory” gradients at anterior temporal/ventral ortbitofrontal effects (Figure 4) extending to the adjacend vmPFC (Figure S5). Nevertheless, given the limited MEG sensor coverage of the vmPFC and a lower resolution of the MEG (vs. MRI) for deeper sources, particularly the observed vmPFC effects require replication using more sensitive modalities.

Second, negative gradients were not only observed in the predicted frontal regions, but also in anterior and posterior temporal brain regions, which have been linked to processes of perception, attention and recognition rather than fear inhibition (see SM2.4.2-5, Figures S2, S6). In line, a systematic review on neural predictive markers for treatment-response identified a consistent contribution of the temporal lobe across studies [39]. Thus, the functional interplay between temporal and frontal structures in treatment-response and generalization should be delineated by future research.

Third, via our variance-analytic approach – opposed to a *genuine predictive* machine learning approach [37] – *group-level associations* between pre-treatment fear generalization and later treatment-response were established. Our supplemental *predictive* approach demonstrates that behavioral and magnetoencephalographic markers of fear generalization might also hold valuable information for the *prediction* of individual exposure outcomes. Yet, prediction models in new, larger and independent samples are needed to estimate the clinical utility of fear generalization.

Noteworthy, we observed no evidence for an influence of the *UCS-type* on behavioral and neural gradient differentiations between responders and non-responders suggesting that general rather than domain-specific fear generalization mechanisms are associated with treatment-responses in spider phobia. This finding seems to contrast with previous evidence for domain-specific conditioning effects in spider phobia [40]. However, while UCS ratings of responding and non-responding patients did not differ here, UCS ratings strongly differed between patients and controls [40]. Thus, the subjective UCS aversiveness should be considered in future studies on the domain-specificity of pathological learning processes in anxiety patients.

To conclude, this study provides initial evidence that pre-treatment differences of behavioral and neural markers of fear generalization are associated with later responses to behavioral exposure. Particularly patients who did not profit from later VRET showed overgeneralized fear responses and aberrant neural generalization effects. Our findings support the relevance of inhibitory learning functions in frontal brain networks during fear generalization and suggest that their spatio-temporal neural reflections underpin the interplay of fear generalization in the laboratory and responses to exposure therapy (ET). These insights may stimulate the development of mechanism-based augmentation strategies for ET. Future investigations on the value of individual fear generalization patterns as predictive markers for treatment outcomes may help to identify anxiety patients who may not profit from ET. This may enable researchers and clinicians to personalize and thereby optimize treatment strategies for this vulnerable patient group [39].

## Data Availability

Data can be provided upon request.

## Acknowledgement

We would like to thank Tilman Coers, Julia Wandschura, Marielle Clerc, Hannah Casper, Sarah Hein, Karin Wilken, Andreas Wollbrink, Ute Trompeter, and Hildegard Deitermann (Institute for Biomagnetism and Biosignalanalysis), Tina Jocham, Jana Scharnagl, and Inge Gröbner (Dept. of Psychiatry, University Hospital of Würzburg); Dominik Grotegerd, Ramona Lennings, Manuel Kraft, Merle Gebauer, Sarah Thissen, Elena Wilkens, Jonathan Repple, Nina Muck, Stella Fingas, Janina Werner, Anna Kraus, and Kordula Vorspohl (Dept. of Psychiatry, University of Münster); Harald Kugel, Jochen Bauer, and Birgit Vahrenkamp (Dept. of Clinical Radiology, University of Münster), Lea Borgmann, Jaqueline Brieke, Aylin Fuchs, Carolin Heinemann, Annika Hense, Valeria Kleinitz, Kaja Loock, Johannes Lücke, and Kathrin Rüb (Institute of Medical Psychology and Systems Neuroscience), for their help and support.

## Statement of Ethics

This research complies with the guidelines for human studies and was conducted ethically in accordance with the World Medical Association Declaration of Helsinki.

## Disclusure Statement

The authors have no conflicts of interest to declare.

## Author Contributions

KR, TS & MJ conceptualized and designed the work. KR, ELJ, JB, FS, HS, MJH, UD & UL conceptualized and implemented the VRET. KR, KH & MJ analyzsed and interpreted the data. KR drafted the work. All authors revised it critically, approved the manuscript to be published and agreed to be accountable for all aspects of the work in ensuring that questions related to the accuracy or integrity of any part of the work are appropriately investigated and resolved.

## Footnotes

1 As behavioral *stimulus-type* by *treatment-response* interactions did not depend on *UCS-type*, this dependency was not tested via permutation analyses on a neural level.

2 Negative ‘Test minus Base’ differences indicate relatively reduced neural responses to stimuli from Test to Base. We here interpret the relative trend observed in these differences.

3 Please find a discussion on the lack of effects in pupil dilation data, collected in a subsample of patients, in the supplementary materials (SM2.3)

