## supplemental materials for "Behavioral and Magnetoencephalographic Correlates of Fear Generalization Are Associated with Responses to Later Virtual Reality Exposure Therapy in Spider Phobia"

#### SM1 Methods

This study was embedded in a prospective longitudinal project employing virtual reality exposure therapy (VRET) as a first-line treatment for specific phobia. It is part of the Transregional Collaborative Research Center (CRC-TRR58) "Fear, Anxiety, Anxiety Disorders" funded by the German Research Foundation. Details on the recruitment pathway (shared by three CRC-TRR58 sub-projects C07, C08, C09), on the highly standardized single-session VRET intervention, and on the a-priori treatment-response criteria have been published [1]. To address the question, whether behavioral and magnetoencephalographic correlates of pre-treatment fear generalization are associated with response to VRET, we here analyzed MEG data collected **before** VRET, as well as clinical outcome measures collected before (Pre-treatment) and after (Post-treatment) the VRET.

##### SM1.1 Participants

All patients were characterized by clinically relevant spider-phobic symptoms indicated by a score of >19 in the German version of the Spider Phobia Questionnaire (SPQ, 2,3).

Patients with a current or lifetime diagnosis of any other comorbid anxiety disorder, obsessive–compulsive disorder, posttraumatic stress disorder, severe major depression, bipolar I disorder, psychotic disorders, substance dependence (except nicotine), self-harming behavior, or acute suicidality were excluded. Exclusion criteria further comprised current (psycho)pharma-cological treatment, current or past psychotherapy with exposure, neurological diseases, pregnancy, and MRI-related exclusion criteria.

Demographic and clinical characteristics of responders and non-responders did not differ before therapy, neither in the behavioral sample (N=89, Table 1, main text), nor in the MEG sample (N=70, Table S1).

[Table: S1]

**SM1.2 Procedure**

Responders and non-responders had a comparable time intervals (in days) between the pre-treatment assessment and the MEG assessment (M=13.03, SD=9.38; t(66.58)=1.54, p=.135), the VRET (M=27.46, SD=10.35; t(87)=1.78, p=.081) and the post-treatment assessment (M=32.70, SD=12.022; t (87)=1.603, p=.113).

**SM1.2.1 Virtual Reality Exposure Therapy (VRET)**

Several days before the one-session VRET treatment (but after the MEG assessment), patients were provided with a detailed psychoeducational manual (adapted from [4]) outlining the function and components of fear, as well as the interplay of cognition, behavior and physiology. The VRET session itself was conducted by board-certified therapists and had a maximum duration of 2.5 hours. The VRET session started with a brief repetition of the manual's key information, followed by an elaboration on the vicious circle of fear in the context of spider phobia, the rationale of behavioral exposure and its mechanism of action. Expectations and apprehensions regarding the VRET and the corresponding experiences *during* the VRET were assessed by means of a pre- and post-treatment protocol obtained before and after the VRET, respectively. The VRET environment was delivered by means of the VR-software (VT+ research systems, VTplus GmbH, Würzburg) and was displayed via an Oculus Rift DK2 head-mounted display. Patients navigated through the virtual environment via a Logitech F310 Gamepad. As described in detail by [1], we selected five standard scenarios, which were ideally completed by each patient. Upon prompts by the therapist, patients verbally rated their individual levels of fear on a scale from 0="no fear at all" to 100="extremely strong fear" before (anticipatory anxiety) and during each scenario. If fear ratings dropped below 20 or stagnated three times in succession, the responsible therapist proceeded with the next scenario. The duration of the behavioral exposure did not differ between responders and non-responders ($t(87)=1.247$, $p=.216$). In the end of the VRET session, the Igroup Presence Questionnaire (IPQ, [5]) was filled in to assess the sense of presence experienced in the virtual environment. Individual scores for each scale (General Presence, Spatial Presence, Involvement and Experience Realism) did not differ between responders and non-responders (all p's >.16).

**SM1.2.2 Behavioral and MEG Assessment on Fear Generalization**

All patients received written and oral information on the generalization paradigm and consented to participate. They then entered the MEG chamber, a sound-attenuated and magnetically shielded room to prevent external magnetic interference. Patients were comfortably seated in the MEG scanner with 90cm distance to the monitor. Prior to the first experimental task, the individual auditory threshold was determined separately for each ear to calibrate the loudness of the US at 60db above individual hearing threshold.

All CS+/CS-/GS stimuli were presented in the center of the screen in front of a grey background (RGB 127, 127, 127; visual angle: 5.1 degrees, edge to edge). All experimental tasks were programmed and presented using the MATLAB-based Psychophysics Toolbox Version 3 ((61); free software, available at www.psychtoolbox.org, Matlab 2016a). The script of the experiment ran via a 2.93GHz Intel Core 2 Duo Processor (4GB RAM) on Microsoft 7 Enterprise, ServicePack 1. The soundcard DMX 6 Fire USB 6-canal Sound-Interface (manufacturer: Fa. Terratec) and a graphic card of the ASUS EAH5750 series (manufacturer: ATI Technologies) were used. Overall, the assessment lasted approximately 2 hours.

**SM1.2.3 Perceptual Midpoint Task**

**Rationale:** Behavioral performance in the PM task, conducted before and after the MEG-assessments, was employed to capture and compare aspects of perceptual discrimination and perceptual learning [6] in responders and non-responders.

**Methods:** The perceptual midpoint (PM) task was adopted from research on perceptual learning mechanisms [6]. Performance in the PM task was employed as an index for different aspects of perceptual discrimination regarding our grating stimuli. The PM task was completed in the beginning (Baseline PM task) and in the end (Test PM task) of each block.

Throughout the PM task, two stimuli (A and B) were positioned in the left and right side of a screen, respectively. One of these stimuli was the CS+ of the respective block, while the other one was the CS-. The allocation to of CS+ and CS- to stimulus A or B was balanced across participants. The PM task comprised three phases (Instruction, Practice and Test) that differed regarding the characteristics of the relevant test stimulus which was presented in the center of the screen between stimulus A and B. In all phases, participants were required to indicate by forced choice button press with the right index or middle finger, whether the test stimulus rather resembled stimulus A (left) or B (right), respectively. In all three phases, stimuli were presented until a key press occurred but at most for a duration of 2000ms. Between each trial a central fixation cross was shown for 500ms.

The instruction (Baseline PM task, Test PM task) and practice phase (Baseline PM task) served to familiarize participants with the task. In these phases, the test stimulus was either stimulus A or stimulus B. Thus, they could easily be matched via a perceptual comparison with stimulus A presented in the left side of the screen and stimulus B presented in the right side of the screen. In the instruction phase each test stimulus was presented three times. Before the subsequent practice phase comprising six additional repetitions of each test stimulus in random order, the experimenter clarified uncertainties remaining after the instruction trials. If more than three errors were made (wrong key, missing response) in the practice block, this block was repeated until the task became clear.

In the subsequent test phase (Baseline PM task, Test PM task), the seven GS of the respective block were presented successively in the center of the screen between stimulus A and stimulus B. The task again required participants to decide by button press whether the presented GS, i.e. stimuli on a perceptual continuum between A and B, perceptually better resembled stimulus A or B. In contrast to the instruction and practice block, this task was more ambiguous, especially for GS ranging in the middle of the perceptual continuum. The test phase consisted of 105 trials in total with each of the seven GSs being presented 15 times in pseudorandomized order.

Regarding the relevant test stimuli, the Baseline PM task, conducted before the Baseline MEG phase, resembled the Test PM task conducted after the Test phase. Note, however, that the *meaning* of stimuli A and B, changed throughout the conditioning and the test phase. Therefore, the comparison of results between the baseline and the test PM task, may inform us on perceptual aspects of fear conditioning and generalization, which may affect the perceptual comparison of the relevant test stimuli (Test PM task: a GS) with the simultaneously presented comparison stimuli A and B (Test PM task: the CS+ and CS-).

**Analysis:** For the PM Task, the relative frequency of classifications as CS+ was determined for each GS, separately for participants and *phases* (Baseline Pm-Task, Test Pm-Task). Weibull cumulative distribution functions ($F(x) = 1 - e^{-(\alpha x)^{\beta}}$) with parameters α (scale parameter) and β (shape parameter) were fitted to these values. The shape parameter β can be interpreted as participants' ability of perceptual discrimination. The point of the Weibull-function with maximal ambiguity of categorization, i.e. the point at which GSs were judged 50% of the time as CS+ and 50% as CS-, is referred to as perceptual midpoint (PM). In addition, a parameter describing the goodness of fit (GoF) of the function to the data was calculated.

Preceding statistical analyses, GoF-parameters (averaged across *UCS-type* blocks) were examined separately for each *phase*. Participants with extremely bad GoF-values, i.e. deviations from the sample´s median by more than four standard deviations, in either *phase* were excluded from further analyses. After exclusion of participants with missing data (N=6) and outliers (N=8) N=76 participants (37 responders and 39 non-responders) entered the final analyses

To investigate differences in β and PM before and after conditioning between responders and non-responders, repeated-measures with the within-subject factor *phase* (Baseline Pm-Task, Test Pm-Task), *UCS-type* (phobia-related, phobia-unrelated), and the between-subject factor *treatment-response* (responder, non-responder) were calculated.

##### SM1.2.4 Recording, Pre-processing, and Analysis of MEG Data

**Recording and Preprocessing:** ERFs were acquired using a 275 sensor whole-head MEG system (Omega 275; CTF, VSM MedTech Ltd., Coquitlam, Canada) with first-order axial gradiometers. Continuous signals in a frequency range between 0 and 150Hz were recorded using a sampling rate of 600Hz. The participants' head shapes were digitized using a 3D tracking device (Polhemus, Colchester, VT, USA). The individual head position in the MEG scanner was tracked by three landmark coils placed on the two ear canals and the nasion.

Pre-processing and further analysis of MEG data was carried out using the MATLAB-based Electromagnetic Encephalography Software EMEGS (Version 3.0, [7]). Continuative statistical analyses were conducted using SPSS Statistics (IBM Corp., Armonk, NY).

Offline, MEG data were filtered using a 48Hz low-pass and a 0.1 high-pass filter and sampled down to 300Hz. Epochs of 800ms duration (i.e., 200ms before to 600ms after stimulus onset) were extracted and baseline-adjusted using the -150ms-0ms baseline interval. Single trials were edited and artefacts were corrected following the method for statistical control of artefacts in high-density EEG/MEG data [8], which (1) detects artifacts in individual sensors, (2) detects global artifacts; (3), replaces artifact-contaminated sensors by spherical spline interpolation that are statistically weighted on the basis of all remaining sensors; and (4) computes the variance of the signal across trials to document the stability of the averaged waveform.

The rejection of artifact-contaminated trials and the interpolation of artifact-contaminated sensors relies on the calculation of statistical parameters for the absolute measured magnetic field amplitudes over time, their standard deviation over time, as well as on the determination of boundaries for each parameter based on their distribution across trials. If the goodness of test topography interpolations based on the residual sensor configuration within a given trial did not reach an a-priori defined minimum criterion (k=0.01 [9]; identical for each subject and run) the respective trial was rejected. If more than 30% of the trials in any run of either the baseline or the test phase did not meet this criterion (e.g. due to continuous movement artifacts or frequent eyeblinks or eye movements across the run), the respective participant was rejected from the MEG-analysis (N=13 participants). A 8x2x2x2 ANOVA with the factors *stimulus-type* (CS+, GS1-7, CS-), *UCS-type* (phobia related, phobia unrelated), *phase* (baseline, test) and *treatment-response* (responder, non-responder) on the number of remaining trials confirmed that the main effect of *treatment-response* and all interactions with *treatment-response* were non-significant (all p's >.19).

**Moving average rationale:** Due to temporal restrictions regarding the patients' vigilance and attention span, each category only included 21 trials (i.e., repetitions per *stimulus-type* in each *phase* and for each *UCS-type*. ERFs to CS+ were merged with those to GS1 (CS+/GS1), those to GS1 were merged with those GS2 (GS1/GS2), etc. This moving average doubled the number of trials per step and thus enhanced the signal to noise ratio by around 40% ($\sqrt{2} \times 100\%$ ), which was specifically important for a reasonable estimation of underlying neural sources. It also reduced high-frequency noise along the vector of *stimulus-type*, while the resolution of the *stimulus-type* gradient function was reduced from 9 to 8 steps only.

**Source reconstruction:** Cortical sources underlying the averaged ERFs were estimated using the L2-Minimum-Norm-Estimates (L2-MNE) method (Hämäläinen & Ilmoniemi, 1994). The L2-MNE is an inverse modelling technique with which distributed neuronal network activity can be estimated. It does not require a priori specifications of the location and/or number of active current dipoles [10]. A spherical shell with 350 evenly distributed dipole pairs (azimuthal and polar direction) with a source shell radius approximately corresponding to the grey matter depth (i.e. 87% of the individually fitted head) was used as source model. The Tikhonov regularization parameter Lambda was set to 0.1. Topographies of source-direction-independent neural activities – the vector length of the estimated source activities at each position – were calculated for each individual participant, condition and time point.

**Non-parametric statistical testing procedure to correct for multiple comparisons** [11]**:** In this procedure, statistical values per time point and estimated dipole entered so-called spatio-temporal cluster masses, if the respective test statistic exceeded a critical alpha level of p=.01 for ANOVAS (categorical treatment response TR-cat) and supplementary regression approaches (dimensional treatment outcomes, TR-dim, for details see SM2.4.4) (sensor-level criteria). Cluster masses within the time-intervals and regions of interest (early and mid-latency IOI: 0-300ms, late: 300-600ms; ROI: anterior; see SM2.4.2 for rationale and effects of supplementary analyses in the posterior ROI) were compared against identical analyses based on 1,000 permuted drawings of the labels of experimental conditions. As ROIs, we used the anterior vs. posterior characterization of the head model implemented in EMEGS (7). This characterization is a rather rough approach separating the model into an anterior vs. posterior half sphere, which covers frontal and anterior temporal regions versus occiptial, parietal and posterior temporal regions respectively. For each permutation, the biggest cluster mass identified within each ROI and IOI was considered. When the cluster mass of the originally labelled conditions was higher than the critical cluster mass of this permutation distribution corresponding to a p-value=.05 (cluster-level criterion), the cluster was considered significant. Temporally or spatially adjacent clusters with qualitatively equivalent results were merged.

**Investigation of baseline differences:** To test for (unexpected) baseline differences of cortical responding of the later Responder and Non-Responder groups before learning we calculated the mean of the estimated neural activity across all experimental conditions (CS+, CS-, GSs) for *UCS-type*-spider and *UCS-type*-face separately as well as across *UCS-type*-spider and *UCS-type*-face at baseline and compared both groups by two sample t-tests. Cluster permutations (NPerm=1,000) of these t-tests with random group labels did not result in any significant clusters with either stronger or reduced neural activity for responders compared to non-responders (for neither $p<.01$ nor $p<.05$ sensor-level significance criteria, all p-cluster > 0.195, for anterior and posterior regions of interest at early (0-300ms) and late (300-600ms) intervals).

##### SM1.2.5 Recording and Pre-processing and Analysis of Pupil Data

For investigation of participants' pupil dilation in response to CS and GS stimuli, the eye tracker EyeLink 1000 Plus (SR Research Ltd., Canada) was used. Pupil data were acquired as external MEG sensor with 600 Hz sampling rate. Due to a software problem, pupil data of 14 of the 70 MEG subjects analyzed here were not stored. Preprocessing and analysis of the pupil data was identical to MEG data analysis but with a temporal dimension only (i.e. just one sensor) and extracted epochs of 2000ms duration.

The analysis of pupil data was performed in parallel with the MEG and behavioral data analysis. Due to strong interindividual differences of mean pupil diameter, Test minus Baseline differences of each participant were normalized by the overall summed root mean square of individual data within the time interval of interest (0-1800ms). Cluster permutation analysis was performed for the complete time interval of interest (0-1800ms) with a first level criterium of $p=0.01$ and a temporal-cluster level criterium of $p=0.05$ (identical to MEG analysis).

##### SM1.2.6. Supplemental Analyses of MEG data: Rationale and Methods

*Linear and Orthogonal Contrasts in Posterior ROI*

In addition to inhibitory gradients peaking at or near the CS-, various studies have revealed positive (excitatory) fear generalization gradients with stronger activity for stimuli approximating the CS+ in wide-spread brain networks including occipito-temporal and parietal brain regions [9, 12, 13]. These posterior brain networks support processes of so-called motivated attention supporting preferential perceptual analyses of emotionally relevant stimuli [14–16]. Electro- and magnetoencephalographic studies have revealed early (< 300ms) [9] and late [9, 17] differentiations of magneto- and electroencephalographic fields and potentials (ERFs/ERPs).

Building upon these findings, we expected late (>300ms) [9, 17] and potentially also early (< 300ms, [9]) positive gradients with stronger activities in response to CS+ compared to GS and CS- in posterior brain regions (occipito-parietal and temporal brain regions). We also explored whether (positive or negative) posterior generalization gradients were associated with responses to treatment.

*Associations between dimensional treatment outcomes and generalization gradients: Effects within identified linear (and orthogonal) clusters*

In supplementary ANCOVAS, we investigated, whether associations between TR-cat and *stimulus-type* in linear and/or quadratic gradients could be replicated with the dimensional treatment outcome measure (TR-dim).

First, parallel to behavioral analyses, we included TR-dim as a covariate, rather than a dichotomized factor on neural activity within clusters displaying linear effects of *stimulus-type* (i.e. independently of TR). Effects are presented in Table S3. Specifically, we investigated associations of percentual SPQ reductions from pre- to post-treatment (i.e. TR-dim) and linear and quadratic polynomial coefficients of generalization gradients within these clusters.

Second, associations between TR-dim and linear polynomial coefficients of generalization gradients were calculated within clusters revealing significant orthogonal linear contrasts (TR-cat, see Table S4). Please note that these results have to be interpreted with caution, given that this analysis is rather circular (clusters were selected based on associations of TR-cat and *stimulus-type*).

*Associations between dimensional treatment outcomes and linear generalization gradients: Cluster permutation analysis*

To generalize our findings from orthogonal analysis comparing linear effects between the group of responders versus non-responders towards a dimensional distribution of treatment outcomes (TR-dim), and to overcome the problem of circularity inherent in analyses testing for TR-dim in clusters revealing orthogonal contrasts, we additionally calculated a polynomial fit of first degree along the dimension of *stimulus-type* (CS- to CS+) and calculated a correlation of TR-dim with the linear polynomial coefficients for each participant, time point and estimated neural source. To control for multiple comparisons a cluster permutation analysis convergent to the categorical analyses was performed (1,000 permutations, p-sensor=0.01, p-cluster=0.05, anterior and posterior ROIs, 0-300ms and 300-600ms IOIs). To identify the strength of correlations and for visualization purposes, Pearson correlation of the respective coefficients within significant clusters (cluster masses) with the individual SPQ reductions were calculated post-hoc. No outliers (>2 scaled median absolute deviation (MAD) above/below the median) were identified in these analyses.

**SM1.2.7 Prediction of individual responses to treatment – an exploratory machine learning approach.**

We performed an additional analysis where we employed machine learning on our behavioral and neural data to predict treatment outcomes on an individual patient level. In such an analysis, the first step is training a predictive model on available data, while the second step is testing the performance of the predictive model on new, previously unused data. If only one dataset is available - as in our study - the available data is split in separate sets for this purpose as long as, crucially, the training process is completely independent from the test data. If this is not the case, and information from the test data “leaks into” the training data, the predictive model may be trained to specifically predict the data in the test set [18]. Therefore, the test set does not represent an independent test of the predictive models performance anymore, and the performance of the predictive model may be considerably overestimated. We therefore decided to conduct a conservative 'naïve' analysis, using the linear and quadratic polynomial coefficients of generalization gradients in six neuronal clusters revealing significant linear contrasts across groups (i.e. independent of therapy response; see Table S3, note that quadratic contrasts revealed no significant clusters on the group level) and the linear and quadratic polynomial coefficients of individual generalization gradients in the behavioral fear- and UCS expectancy rating. To estimate polynomial coefficients we averaged responses across the two *UCS-types*.

We used scikit-learn 0.23.2 [19] for machine learning. As our total sample size was limited but the leave-one-out cross-validation (CV) framework has been demonstrated to be unstable [20], we applied a 5-fold CV approach which has been suggested in such cases by Scheinost and coworkers [21]. Classification was done via Random Forests, and hyperparameters were tuned within nested 5-fold CVs with 100 iterations within the train sets. The means of balanced accuracy, sensitivity, specificity and area-under-the-curve (AUC) over the five folds were used as performance metrics. Finally, we used a permutation test with 5000 iterations to evaluate whether each predictive model was superior to chance level [22]. To this end, in each iteration labels in the train and test folds were randomly shuffled and the mean balanced accuracy across the five folds with shuffled labels was compared to the real mean balanced accuracy. The p value was then calculated as $\sum((\text{accuracy}_{\text{test}} < \text{accuracy}_{\text{permutation}}) + 1) / (n_{\text{permutations}} + 1)$.

Note that this 'naïve' analysis most probably underestimates the true performance of a fully predictive model, since we here refrained from sophisticated feature reduction and extraction methods to circumvent biases based on the previously mentioned knowledge about the dataset.

### SM2 Results and Discussion

#### SM2.1 Clinical effect of VRET

VRET effectiveness was also reflected in the secondary outcome measures: Behavioral avoidance (BAT final distance) decreased from pre- to post treatment assessement (t(88)=11.478, p<.001, d=1.22; $M_{Pre}$ = 172.80cm, $SD_{Pre}$ = 75.55cm; $M_{Post}$ = 88.72cm, $SD_{Post}$ = 69.38cm). Likewise, the CGI revealed a reduction of clinical severity from pre- to post treatment assessment (Wilcoxon-Test: N=86, Z=7.183, p<.001; not ill: $N_{Pre}$ = 0, $N_{Post}$ = 3; marginally ill: $N_{Pre}$ = 0, $N_{Post}$ = 7; mildly ill: $N_{Pre}$ = 10, $N_{Post}$ = 44; moderately ill: $N_{Pre}$ = 42, $N_{Post}$ = 39; markedly ill: $N_{Pre}$ = 43, $N_{Post}$ = 4; severely ill: $N_{Pre}$ = 1, $N_{Post}$ = 0). Note that percentual SPQ and BAT reductions from pre to post-assessment were moderately correlated only (r(88)=.372, $r^2$=0.138 p<.001). Evidence for longterm-effects in primary and secondary outcomes is presented in Leehr, Roesmann et al. (subm) [23].

#### SM2.2 Behavioral Data

##### SM2.2.1 Fear Ratings After Conditioning

*Fear Ratings of CS+ and CS- (N=89 participants)*

After conditioning, we expectedly found a main effect of the factor *stimulus-type* with higher fear ratings for CS+ compared to CS- (F1,87)=163.166, p<.001, $\eta^2$=0.652), a main effect for *UCS-type* (F1,87)=58.871, p<.001, $\eta^2$=0.404) with higher fear ratings for both CS in the phobia-related blocks and an interaction of *stimulus-type* and *UCS-type* (F(1,87)=28.900, p<.001, $\eta^2$=0.249), with a stronger CS+/CS- differentiation in the phobia-related block (F1,87)=138.729, p<.001, $\eta^2$=0.615) than in the phobia-unrelated block (F1,87)=45.981, p<.001, $\eta^2$=0.346). Neither of these effects was modulated by *treatment-response* (*stimulus-type* by *treatment-response* (F(1,87)=0.001, p=.980, $\eta^2$=0.000; *UCS-type* by *treatment-response* (F(1,87)=2.684, p=.105, $\eta^2$=0.030; *stimulus-type* by *UCS-type* by *treatment-response* (F(1,87)=0.892, p=.347, $\eta^2$=0.010).

*Fear Ratings of UCS*

Fear ratings of UCS obtained after the fear conditioning procedure (see Figure 2, bottom, main text) indicated higher fear levels in response to phobia-related compared to phobia un-related audiovisual UCS (F(1,87)=119.21, p<.001, $\eta^2$=.578). However, this main effect of the factor *UCS-type* was not modulated by the *treatment-response* (F(1,87)=0.009, p=.927, $\eta^2$=0.000). Average fear ratings of UCS did not differ between responders and non-responders (F1,87)=0.338, p=.562, $\eta^2$=0.004).

##### SM2.2.2 Fear Ratings and UCS Expectancy Ratings after Test Phase

*Fear Ratings (N=89 participants, additional effects):*

In addition to the effects of interest, reported in the main text, we observed a main effect of *UCS-type*, with overall higher fear ratings for all stimuli presented in the blocks with the phobia-relevant UCS (F(1,87)=39.143, p<.001, $\eta^2$=.310). This effect was not associated with the factor *treatment-response* (F(1,87)=2.615, p<.110, $\eta^2$=.029). However, there was a significant interaction of *stimulus-type* and *UCS-type* (linear gradient: F(1,87)=19.963, p<.001, $\eta^2$=.187; quadratic gradient: F(1, 87)=3.173, p=.078, $\eta^2$=.035). Fear ratings obtained in the phobia-related block revealed shallower linear gradients (linear: F(1,87)=60.122, p<.001, $\eta^2$=0.409, quadratic: F(1,87)=0.018, p=.893, $\eta^2$=0.000) than those in the phobia-unrelated block (linear: F(1,87)=135.654, p<.001, $\eta^2$=0.609, quadratic: F(1,87)=5.761, p=.019, $\eta^2$=0.062). The main effect *treatment-response* was not significant (F(1,87)=0.197, p=.658, $\eta^2$=.002).

*UCS Expectancy Ratings (N=89 participants, additional effects):*

In contrast to the fear ratings, the main effect of *UCS-type* was not significant (F(1,87)=2.259, p=.136, $\eta^2$=.025). The interaction *UCS-type* by *treatment-response* (F(1,87)=0.883, p=.350, $\eta^2$=.010) and the interaction *stimulus-type* by *UCS-type* were also both non-significant (linear gradient: F(1,87)=0.033, p=.857, $\eta^2$=.000; quadratic gradient: F(1, 87)=127, p=.723, $\eta^2$=.001). The main effect *treatment-response* was not significant (F(1,87)=0.008, p=.927, $\eta^2$=.000).

*Fear Ratings and UCS-Expectancy Ratings (N=70 participants, MEG sample)*

For the MEG sample, effects of fear ratings and UCS-expectancy ratings after the test phase were qualitatively identical to the sample of 89 partipants and are presented in Table S2

[Table S2]

**SM2.2.3: Perceptual midpoint task**

76 participants entered the analysis. The shape parameter β, an index for participants' ability of perceptual discrimination, was not modulated by *phase* ($F(1,74)=0.741$, $p=.600$, $\eta^2=0.004$). No interactions of *treatment-response* and *phase* ($F(1,74)=0.741$, $p=.392$, $\eta^2=0.010$) or of *treatment-response* by *phase* by *UCS-type* ($F(1,74)=0.548$, $p=.462$, $\eta^2=0.007$) were observed. Interactions of *phase* by *UCS-type* were also non-significant ($F(1, 74)=1.368$, $p=.246$, $\eta^2=0.018$). Thus, perceptual discrimination abilities were not affected by the conditioning procedure.

The main effect of *treatment-response* ($F(1,74)=2.848$, $p=.096$, $\eta^2=0.037$) and the interaction of *treatment-response* and *UCS-type* ($F(1,74)=3.441$, $p=.068$, $\eta^2=0.044$) were also non-significant.

The modulated perceptual midpoint was not modulated by *phase* ($F(1,74)=0.162$, $p=.688$, $\eta^2=0.002$. No interactions of *treatment-response* by *phase* ($F(1,74)=0.295$, $p=.588$, $\eta^2=0.004$) or of *treatment-response* by *phase* by *UCS-type* ($F(1,74)=0.177$, $p=.675$, $\eta^2=0.002$) were observed. Interactions of *phase* by *UCS-type* were also non-significant ($F(1, 74)=0.058$, $p=.810$, $\eta^2=001$). Thus, the perceptual midpoint was not affected by the conditioning procedure.

All other effects were non-significant (*treatment-response*: $F(1,74)=2.073$, $p=.154$, $\eta^2=027$; *UCS-type*: $F (1, 74)=0.314$, $p=.577$, $\eta^2=0.004$, *treatment-response* by *UCS-type*: ($F (1, 74)=0.807$, $p=.372$, $\eta^2=0.011$)

**Discussion:** The perceptual midpoint task yielded no evidence for learning-dependent modulations of discrimination abilities (shape parameter β) or perceived perceptual midpoints in the PM task, at contrast with previous research [6].

**SM2.3 Pupil Data**

*Results:*

Linear positive contrast (CS- < GS1 < GS2 < GS3 < GS4 < GS 5 < GS6 < GS7 < CS+) analysis across the dimension of *stimulus-type* revealed a significant (p-cluster<.001, $F(1,54)=34.174$, $p<.001$, $\eta^2=0.388$) temporal cluster in a time interval between 1063 and 1800ms after stimulus onset (see Figure S1). Post-hoc analysis of this cluster revealed that neither linear nor quadratic gradients differed between responders and non-responders (linear: $F(1,54)=0.078$, $p=.781$, $\eta^2=.001$; quadratic: $F(1,54)=0.309$, $p=.581$, $\eta^2=.006$). The three-way interactions *stimulus-type* by *treatment-response* by *UCS-type* were non-significant for linear ($F(1,54)= 3.244$, $p=.077$, $\eta^2=.057$) and for quadratic trends ($F(1,54)=0.336$, $p=.564$, $\eta^2=0.006$). The main effects of *treatment-response* and *UCS-type* as well as the interactions *UCS-type* by *stimulus-type* and *UCS-type* by *treatment-response* were all non-significant (all Fs<1). Negative linear contrast and orthogonal linear contrast analyses testing for differential linear effects of *treatment-response* did not reveal any significant effects.

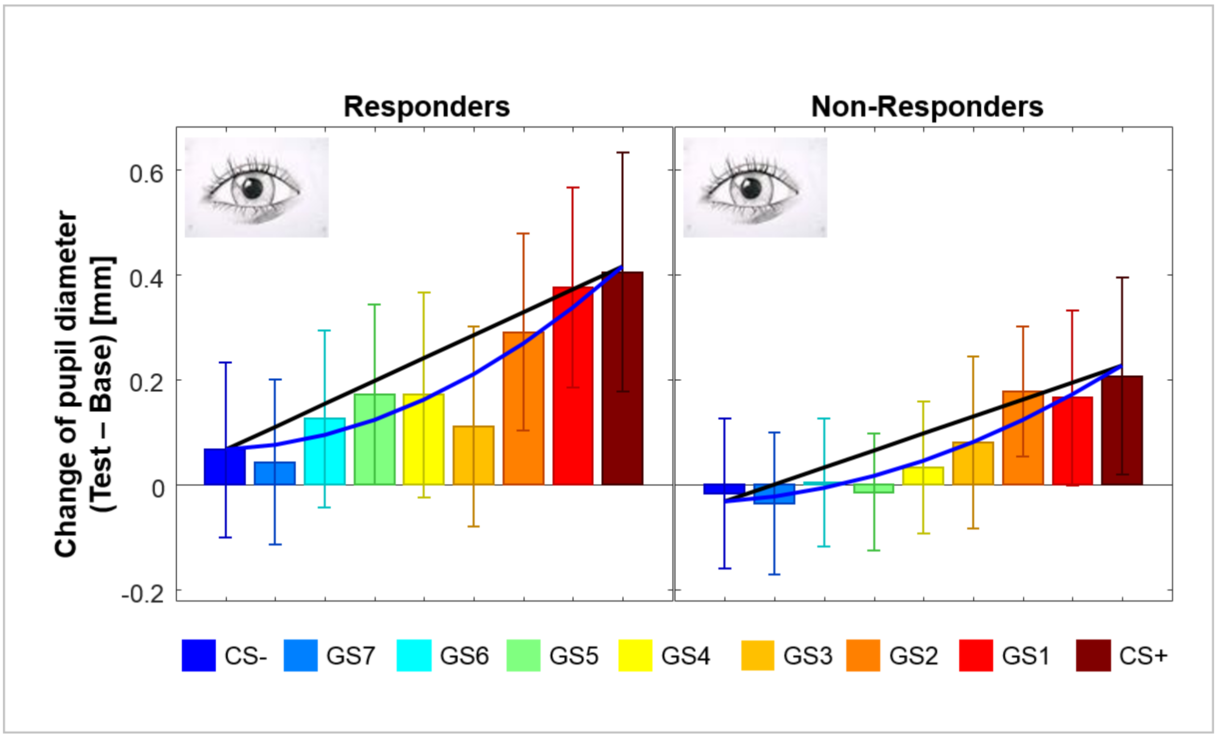


**Figure S1:** Psychophysiological generalization effects indicated by the change of pupil diameter (Test minus Base, 1063-1800ms after stimulus onset, N=56). Responders and non-responders revealed similar effects. Error bars denote 95% confidence intervals.

*Discussion:*

Pupil dilations were employed as a robust psychophysiological readout of fear learning [24, 25] and mirrored linear generalization effects observed in fear- and UCS-expectancy ratings. However, neither linear nor quadratic contrasts in pupil dilations significantly differed between responders and non-responders[1]. This lack of effects might be related to several aspects, including methodological ones: First, following our primary hypotheses, our paradigm was optimized for the collection of MEG data: SOAs (~800ms) and ISIs (1850+/-300ms) were kept relatively short, to keep the experiment within reasonable time limits, and profit from an optimal number of trials to enhance the signal to noise ratio allowing a reasonable estimation of neural sources. For the detection of pupil responses, this setup is not optimal: SOAs (and ISIs) in

[1] Yet, a marginally significant three-way interaction of *stimulus-type* by *treatment-response* by *UCS-type* was observed for linear gradients, suggesting some influence of the *UCS-type* on pupil responses. Preliminary exploratory analyses of pupil data further suggest complex (non-linear and non-quadratic) interactions in pupil data that warrant further investigation.

conditioning studies focusing on pupil dilation (e.g. [24, 25]), are usually much longer (> 2 seconds). As consequence of short SOAs, we found group-independent linear generalization effects (1063-1800ms after stimulus onset) after offset of CS+ and CS-. Thus, potential group differences might have been covered by offset effects.

Second, we obtained pupil data only in a subsample of patients, which limits the statistical power of potential interaction effects in this outcome measure. Thus, despite the lack of effects here, generalization effects in pupil responses might be informative regarding response prediction in an optimized experimental setting for the collection of pupil responses.

#### SM2.4 MEG data

##### SM2.4.1 Effects in anterior networks

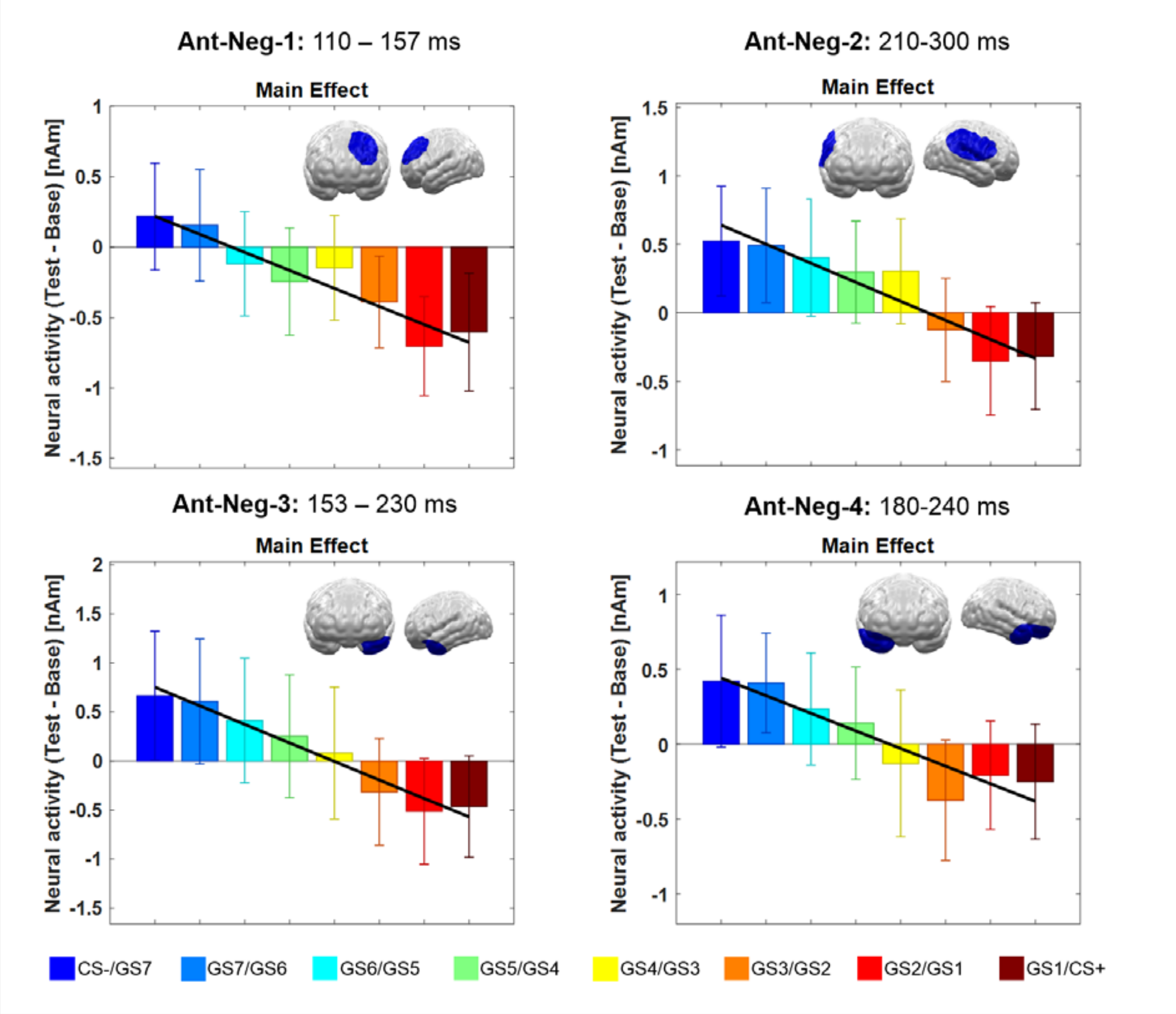

**Figure S2:** Spatiotemporal clusters revealed by the the permutation test of negative (neg) linear contrasts (indicated in blue) in the anterior (Ant) ROI. Significant negative ("inhibitory") gradients were observed in the early time interval (< 300ms) only. Bar graphs show the regional neural activity in the displayed clusters. Error bars denote 95% confidence intervals.

*Negative linear contasts*

A cluster in left dorsolateral prefrontal regions (see Figure S2, Ant-Neg-1) yielded significance as early as 110-157ms after stimulus onset. Details on this cluster are described in the main text, as contrasts differed between responders and non-responders.

An additional cluster revealing a negative inhibitory response was observed between 257 and 300ms in right ventrolateral prefrontal regions (p-cluster=.004). As this cluster was localized spatio-temporally adjacent to a cluster in the right TPJ in the posterior ROI (210–260ms, p-cluster=.011), we merged these two clusters. The merged cluster (210-300ms, see Figure S2, Ant-Neg-2) revealed a significant inhibitory linear gradient with stronger activations for the CS- compared to the CS+ ($F(1,68)=28.177$, $p<.001$, $\eta^2=.293$). Responders showed overall stronger activations in this cluster ($F(1,68)=7.219$, $p=.009$, $\eta^2=.096$), but gradients did not significantly differ between responders and non-responders ($F<1$).

Three clusters in anterior regions spanning the bilateral temporal poles (left: 153–230ms, p-cluster=.002, see see Figure S2, Ant-Neg-3); right 180–220ms, p-cluster=.01) and extending to right ventral frontal brain regions (217-240ms; p-cluster=.0041) revealed negative inhibitory gradients. No modulatory effects of the factor *treatment-response* were observed for these clusters. Due to the temporal and spatial vicinity of clusters in right anterior temporal and ventral orbitofrontal regions and qualitatively identical effects, we merged these clusters for visualization purposes. The merged cluster (180-240ms, see Figure S2, Ant-Neg-4) revealed a significant inhibitory linear gradient ($F(1,68)=16.524$, $p<.001$, $\eta^2=.195$), which again was not affected by *treatment-response*.

**SM2.4.2 Supplemental Analyses I: Effects in posterior networks**

*Confirmatory Analysis: Positive linear contrasts*

The predicted positive gradients in posterior brain networks were found at late (>300ms) latencies only. The cluster-based permutation test for the positive linear contrast revealed two significant clusters (437–490ms, p-cluster=.011; 527–567ms, p-cluster=.01). Both clusters were located in overlapping centro-parietal brain regions and were merged for visualization purposes (see Figure S3 A, Post-Pos-1). Visual inspection of the statistical maps for the positive linear contrast in this merged cluster (Figure S3 A, top right), suggested that these effects extended towards right occipital networks. However, only centro-parietal parts of these effects yielded significance on both the sensor- and cluster-level. In addition to the linear positive gradient, the earlier cluster (437–490ms) yielded a significant main effect of the factor *UCS-type* ($F(1,68)=4.851$, $p=.031$, $\eta^2=.067$) with overall stronger differential neural activity in response to CS+, CS- and GS in the phobia related compared to the phobia unrelated block, independently of the factor *stimulus-type*.

*Exploratory Analysis: Orthogonal linear contrast*

This analysis revealed a significant orthogonal contrast in right temporo-occipital regions at early latencies (130–160ms, p-cluster=.048, Figure S3 B, Post-Int-1). This effect was characterized by a positive excitatory gradient (CS- < GS < CS+, $F(1,35)=6.472$, $p=.016$, $\eta^2=.156$) in the group of responders, while non responders showed the opposite pattern: CS- > GS > CS+; $F(1,33)=7.435$, $p=.010$, $\eta^2=.184$)[2]. Again, treatment-response-dependent linear gradients within these clusters were not modulated by *UCS-type* ($F(1,68)=0.003$, $p=.956$, $\eta^2=.000$).

[2] The negative Test minus Base differences indicate habituation effects for all stimuli from Test to Base, which were relatively weaker (less negative) for CS+ compared to CS- processing in the group of responders while the group of non-responders revealed the opposite CS+ vs CS- habituation effect.

*Exploratory Analysis: Negative linear contrasts*

Surprisingly, analyses in the early IOI (posterior ROI) yielded two clusters, that revealed negative instead of the predicted positive gradients. One of these clusters was merged with an anterior cluster, due to its temporal and special vicinity (for details see above). The other cluster was significant from 47 to 100ms after stimulus onset in right temporal brain regions (p-cluster=.017, see Figure S3 C, Post-Neg-1). Interestingly, this cluster not only revealed a significant linear negative gradient, but also a significant interaction of *treatment-response* by *stimulus-type*, indicating that linear gradients differed across groups ($F(1,68)=4.057$, $p=.048$, $\eta^2=.056$). Linear contrasts conducted separately for responders and for non-responders indicated significant linear effects in both groups (responders: $F(1,35)=6.114$, $p=.018$, $\eta^2=.149$; non-responders: $F(1,33)=24.833$, $p=.001$, $\eta^2=.429$). Treatment-response-dependent linear gradients within these clusters were again not modulated by the *UCS-type* ($F(1, 68)=0.375$, $p=.542$, $\eta^2=.005$).

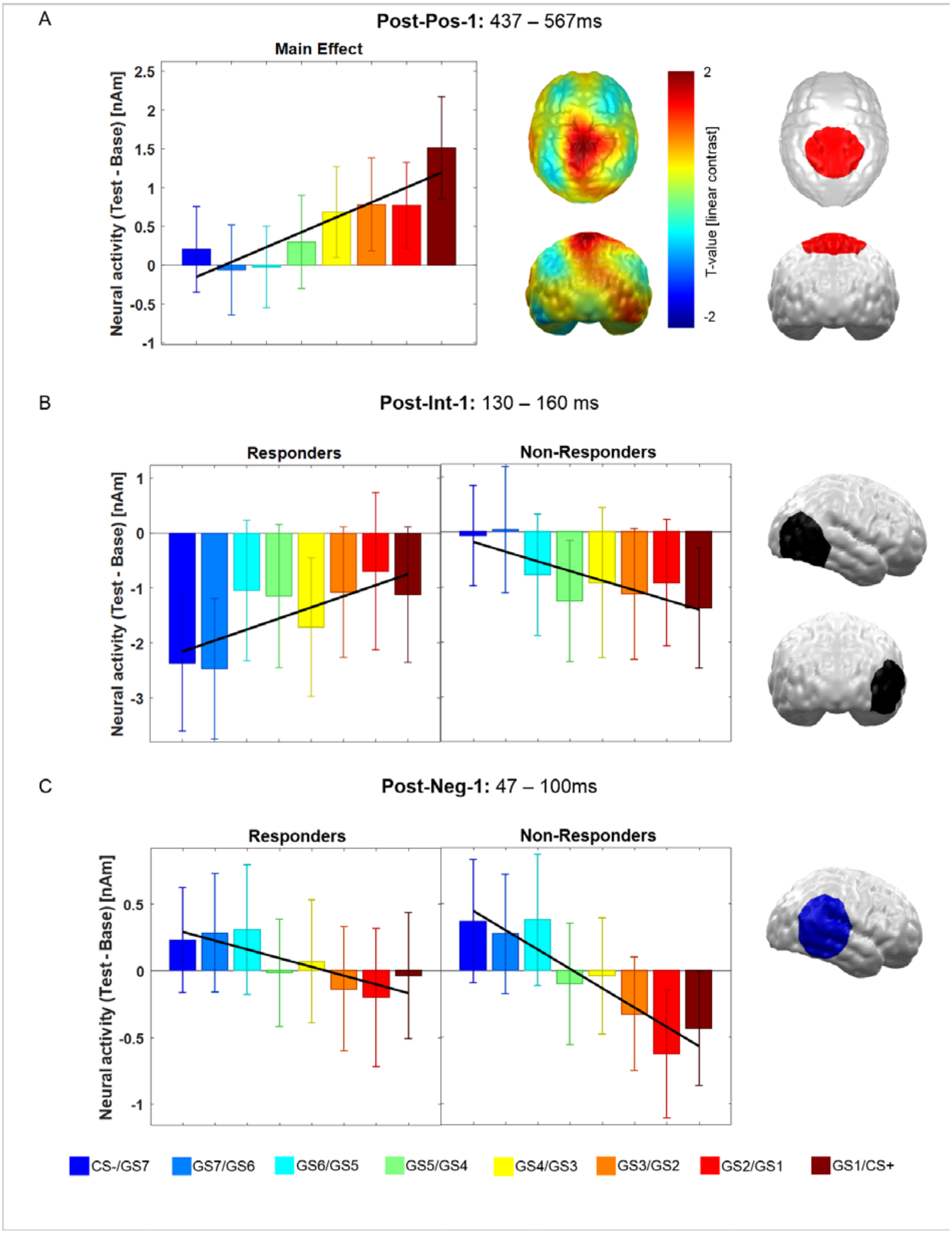


**Figure S3:** Significant spatiotemporal clusters showing generalization gradients as revealed by permutation tests conducted in the posterior region of interest. Clusters revealing significant positive (Pos) gradients are visualized in red. Clusters revealing interaction effects (Int) in linear gradients, i.e. orthogonal contrast, are displayed in black. Clusters revealing significant negative gradients (Neg) are displayed in blue. Bar graphs show the regional neural activity in the displayed clusters. Error bars denote 95% confidence intervals.

A) The predicted positive (Pos) linear main effects with stronger activation for the threat-signaling CS+ compared to the CS- were revealed for the late time interval only (> 300ms). The distribution of t-values for this contrast (right) suggests that the effect extended from centroparietal regions (see also cluster Post-Pos-1, in red) to occipital areas.
B) Significant orthogonal clusters (Interaction, Int) in posterior networks were observed in the early time interval (< 300ms). Excitatory linear gradients were observed for responders only, while non-responders revealed the opposite pattern.
C) Additional exploratory analyses testing for negative (Neg) gradients in the posterior ROI yielded a significant cluster at early latencies (47-100ms). This cluster was further modulated by the factor treatment response.

##### SM2.4.3: Supplemental Analyses 2: Associations between dimensional treatment outcomes and generalization gradients: Effects within identified linear (and orthogonal) clusters

*Results:*

[Table: S3]

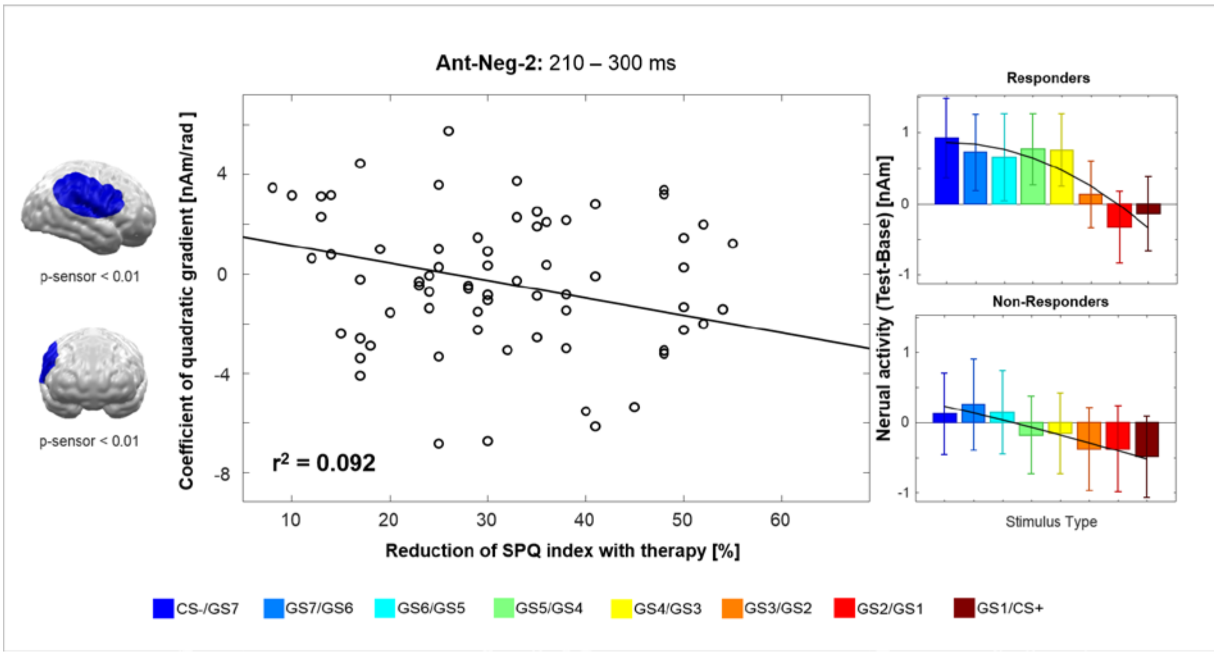


**Figure S4:** One spatiotemporal cluster revealed by the the permutation test of negative (neg) linear contrasts (indicated in blue) in the anterior (Ant) ROI (Ant-Neg-2, see also Figure S2) revealed significant associations of dimensional treatment outcomes (TR-dim) as indexed by percentual reductions of the SPQ from the clinical pre- to post-treatment assessement and coeffeicients of quadratic gradients in STIMULUS TYPE. For visualization purposes only, quadratic gradients are visualized separately for responders and non-responders according to the categorical outcome (TR-cat) in bar-charts in the right panel of the Figure.

[Table: S4]

<u>Discussion:</u>

We found evidence for an interaction of quadratic gradients and TR-dim in a cluster revealing linear effects of *stimulus-type* in right ventrolateral prefrontal regions extending to the right TPJ (Cluster Ant-Neg-2, Figure S4). This cluster did not reveal interaction effects when considering TR-cat only. In accordance with the hypothesis, that VLPFC regions are involved in fear inhibition, this cluster indicates that better responses to treatments are associated with relatively stronger brain activations to CS- and CS- like GS vs. CS+ stimuli. This effect is particularly interesting, as it mirrors the quadratic effect in fear-ratings observed on a behavioral level.

##### SM2.4.4: Supplemental Analyses 3: Associations between dimensional treatment outcomes and linear generalization gradients - Cluster permutation analysis

<u>Results:</u>

An anterior cluster covering vmPFC/rOFC and anterior temporal regions got significant (p-cluster = .008) in a late time interval between 427 and 497 ms after stimulus onset (see Figure S5). A reduction of the first level significance criterion from p-sensor = .01 to p-sensor= .05 revealed that this effect was not strictly right lateralized but also covered more medial vmPFC regions. The post-hoc analysis revealed a negative correlation ($r(68)=-0.488$; $r^2=0.238$; $p<.00001$) with a large effect size [26] in this cluster with increasingly steeper negative linear gradients with improving treatment outcome (i.e. percentual SPQ reduction). In line with orthogonal contrasts in the TR-cat analyses (see Figure 4B, Ant-Int-2), responders revealed a stronger differentiation of safety and threat processing compared to non-responders in vmPFC/OFC regions spanning anterior temporal regions. To allow a comparison with effects identified by the categorical analysis, we here also visualized the effects separated by the a-priori defined response groups. While neural activity in dlPFC/vlPFC clusters indentified via negative linear and orthogonal contrasts (Figure 4, Ant-Neg-1, Ant-Int-1) yielded significant associations with TR-dim, they failed to reach significance in this cluster permuation analysis.

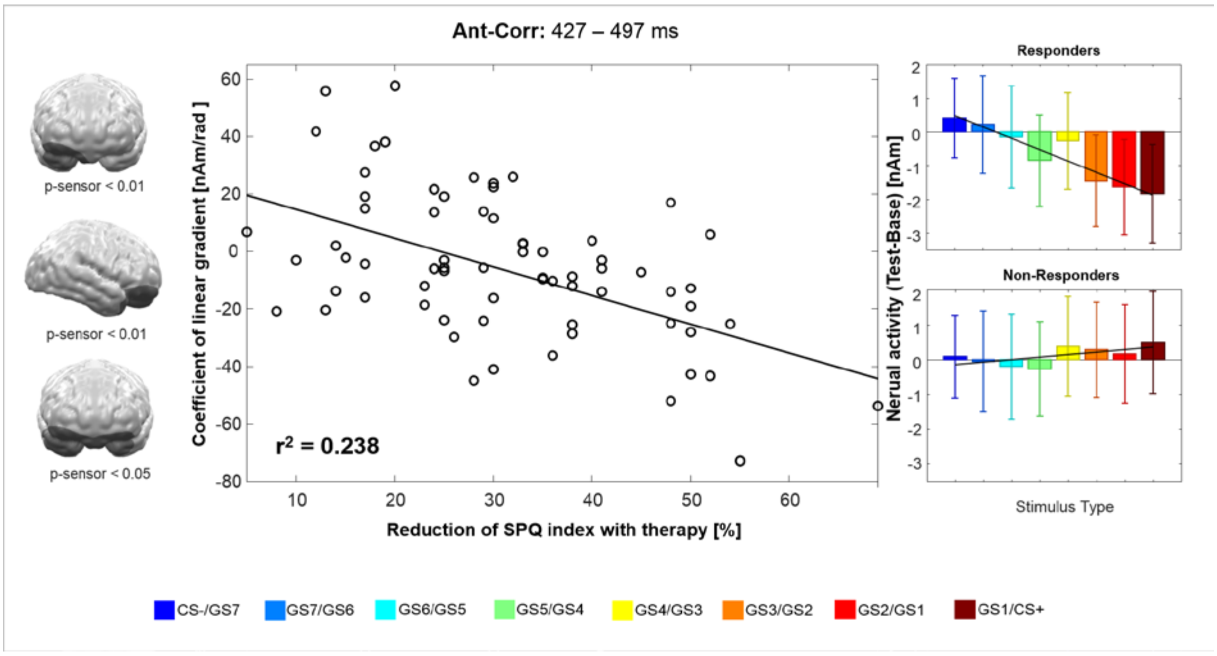


**Figure S5:** Spatiotemporal cluster showing a significant association of the linear polynomial parameter and dimensional treatment outcomes (TR-dim) as indexed by percentual reductions of the SPQ from the clinical pre- to post-treatment assessement in the anterior ROI. When lowering the original sensor-level significance criterion from .01 to .05, the cluster covered the bilateral anterior temporal pole as well as the vmPFC. For visualization purposes, linear gradients are visualized separately for responders and non-responders according to the categorical outcome (TR-cat) in bar-charts in the right panel of the Figure. Steeper negative gradients were associated with better treatment outcome.

Additionally, a posterior cluster at left occipito-temporal regions got significant (p-cluster = 0.022) in an early time interval between 47 and 110 ms after stimulus onset (see Figure S6). The post-hoc evaluation revealed a positive correlation ($r(68)=0.575$; $r^2=0.331$; $p<0.00001$) of large effect size [26] in this cluster with increasingly steeper positive gradients with improving response to therapy. A reduction of the first level significance criterion from p-sensor = .01 to p-sensor= .05 revealed that this effect was not strictly left lateralized but occurred bilaterally. Thus, as for the right occipito-temporal regions in the identical (see Figure S3 C) and a slightly later time interval (see Figure S3 B), responders revealed a more positive gradient with relatively increased threat compared to safety signal processing while non-responders showed the opposite effect. Interestingly, within this early time interval (47-110 ms) the anterior cluster at vmPFC/rOFC regions which was identified at later time intervals (427-497 ms; Figure S5), also revealed a significant negative correlation with therapy outcome (r(68)=-0.239; $r^2$=0.057; p<0.05) indicating a rather sustained differential effect of prefrontal inhibition on sensory processing with quite early onset.

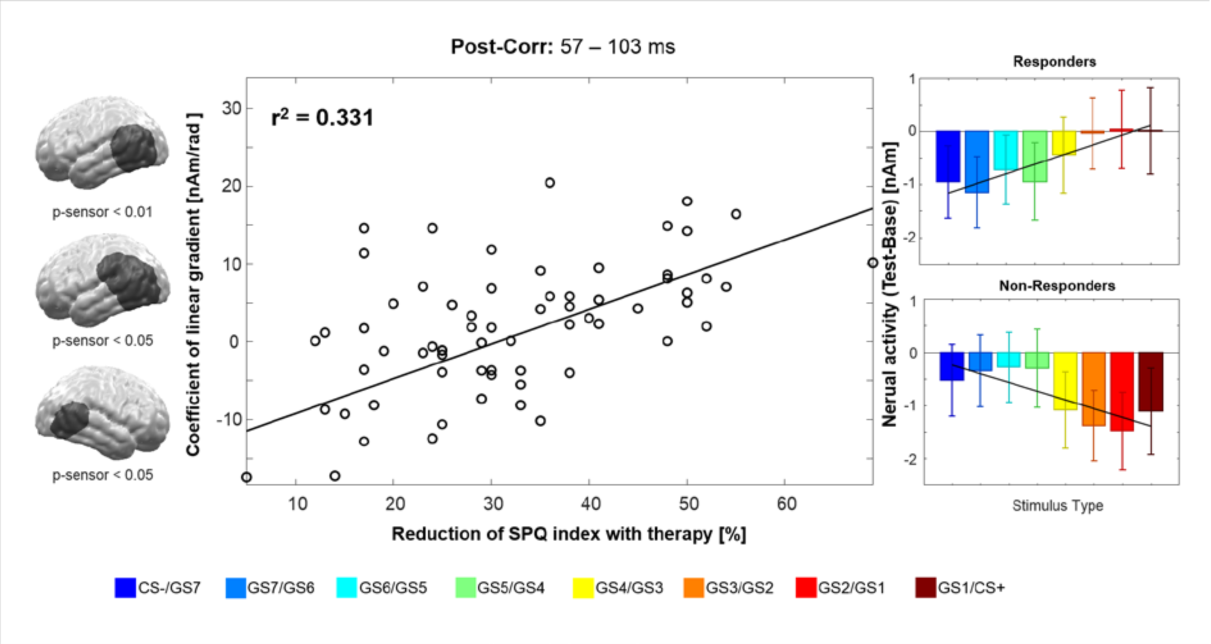


**Figure S6:** Spatiotemporal cluster showing a significant association of the linear polynomial parameter and dimensional treatment outcomes (TR-dim) as indexed by percentual reductions of the SPQ from the clinical pre- to post-treatment assessement in the posterior ROI. When lowering the original sensor-level significance criterion from .01 to .05, bilateral occipito-temporal clusters yielded significance. For visualization purposes, linear gradients are additionally visualized separately for responders and non-responders according to the categorical outcome (TR-cat) in bar-charts in the right panel of the Figure. Relatively more positive gradients were associated with better treatment response.

#### *Discussion:*

Overall, results of different conceptualizations of treatment outcome (TR-cat, main text; and TR-dim, supplementary material) qualitatively converge in the notion of associations between anterior (negative) gradients and treatment response. They suggest that effects observed in anterior temporal/ventral orbitofrontal regions extended to (bilateral) vmPFC regions. This finding nicely fits with previous research that led us to predict vmPFC effects [12, 27–29], which could not be shown via orthogonal contrasts in responders vs. non-responders. In combination with effects revealed in posterior brain regions, these findings substantiate the idea that the strength of inverse relationships between anterior inhibitory negative gradients and positive gradients in the visual pathway might predict therapeutic success. Such network-based hypotheses should be targeted by future research.

Overall additionial effects resulting from analyses on dimensional outcomes fit well into the overall picture that aberrant fear-inhibitory processes in non-responders compared to responders are mediated by dorso-and ventrolateral prefrontal brain regions as well as vmPFC brain regions.

**SM2.4.5 Overall discussion of effects in posterior brain regions**

As predicted, our study revealed excitatory gradients peaking at the threat-signaling CS+ in the posterior region of interest (see Figure S3 A, for responders: see also Figure S3 B and Figure S6). Excitatory gradients in wide-spread brain networks including occipito-temporal (below significance here) and parietal brain regions are in line with previous research [9, 12, 13]. However compared to previous studies [9, 12], the observed effects peaked in more superior dorsal parts of the parietal cortex. One likely explanation for differences in the observed topographies refers to the characteristics of the employed stimuli. Previous studies employed differently sized rings [12] or faces [9] as generalization stimuli. The discrimination of these stimuli might be supported by inferior parietal and temporal processes. Here, by contrast, participants needed to particularly attend to and discriminate stimuli with different orientations. These functions are supported by dorsal parietal networks, including the superior parietal lobule [30], in which we found generalization effects at later intervals (> 300ms).

Electro-and magnetoencephalographic evidence for temporal aspects of motivated attention in fear generalization are sparse. However, in line with the current study, two previous ERP/ERF studies on healthy subjects have revealed evidence for late (> 300ms) generalization effects [9, 17]. In the study by Nelson et al. [17], these late generalization effects were reflected in the so-called late positive potential (LPP). The LPP, an emotion-sensitive electro-physiological component, has been associated with late attentional processes in response to motivationally relevant stimuli [31] that are subject to compensatory strategies like avoidance [32] and cognitive biases such as individual levels of intolerance of uncertainty [17]. Thus, contrary to our findings, one might have expected associations between pre-treatment generalization effects in this component and later treatment responses.

Instead, we found treatment-response-dependent gradients in earlier time intervals (< 300ms, see Figure S2 B). Between 130ms and 160 after stimulus presentation, we found evidence for differential pre-treatment generalization gradients in a right temporal cluster. This finding again converges with evidence for a predictive role the temporal cortex for later treatment response and supports previous speculations that "neural substrates of visual object processing and recognition may hold predictive potential for treatment outcome" ([33], p.158). Specifcally, we observed a positive gradient peaking at the CS+ only in the group of treatment responders. By contrast, positive generalization effects at early latencies, as previously also observed for healthy subjects [9], could not be replicated in non-responders, who showed the opposite pattern. A key component reflecting a prioritized perceptual analysis of emotionally relevant compared to neutral stimuli in visual brain regions between around 130 and 300ms is the early posterior negativity (EPN) [31, 34, 35]. This component is regarded an index of motivated attention [14–16, 31]. Following functional interpretations of EPN effects in previous studies claiming preferential perceptual analyses of emotionally relevant compared to neural stimuli [31], we infer that responders compared to non-responders analyze threat-signaling stimuli in a prioritized manner.

One might speculate that this analysis *drives*/*enables* the maintenance of inhibitory processes in frontal networks (see main text), which may ultimately result in less overgeneralization on a behavioral level. On the other hand, there is also compelling evidence that signals from areas outside the visual cortex (e.g. frontal structures) may drive plastic changes in sensory sensitivity during fear conditioning via feedback loops [13, 36]. Our analyses cannot establish causal or directional links between effects observed in frontal and posterior networks. Future work should address this important question, for example by combining generalization paradigms with targeted brain stimulation (e.g. [37, 38]).

Unexpectedly, we found one very early cluster in right temporal regions (47–100ms) showing negative gradients peaking at the safety signaling CS-. These results are at contrast with our previous study revealing opposite effects with stronger activity peaking at stimuli similar to the CS+ at similar latencies [9]. These differences in findings might be caused by several factors like different samples (spider phobic patients vs. healthy participants), stimulus characteristics (gratings vs. faces), differences in the paradigm (distinct awareness vs no contingency-awareness on the dimension of generalization). Interestingly, however, these very early clusters (once more) suggest pre-treatment differences in fear generalization in brain regions supporting perceptual processes [33]. Complementing this, previous studies on fear generalization have linked cortical and subcortical structures with processes of emotional attention [9], perception [29], as well as pattern separation, recognition and the modulation of fear responses (e.g. hippocampus [12, 39]). To conclude, the observed pre-treatment differences between responders and non-responders in posterior networks in combination with evidence for differences in frontal inhibitory gradients provide initial evidence that a complex spatiotemporal interplay of both fear inhibitory and sensory processes may mediate behavioral levels of fear generalization. Importantly, we provide first evidence that pre-treatment differences in this interplay may act as moderators of later responses to behavioral exposure.

##### SM2.4.6 Limitations and future directions

Future neuroscientific studies should investigate the functional role of generalization gradients in more depth, for example by associating them more directly with behavioral measures of fear inhibition or recognition. Additionally, inverse MEG source modeling of subcortical structures might provide important information about subcortical affective learning and generalization mechnisms and their crosstalk with cortical regions. Because of our hardware (MEG radial gradiometer system, which additionally reduces resolution of subcortical structures) and due to missing a priori spatio-temporal information about potentially simultaneously active subcortical and cortical sources, we here opted to apply an L2-Minimum-Norm based on a head model excluding subcortical structures. We acknowledge, that neural activity of deeper structures that were not covered by the source model might project their activity to modelled sources at the cortical surface leading to a limited spatial accuracy. However, here we opted to minimize the chances for false negative findings in cortical structures at cost of information about subcortical activity and spatial accuracy. Future research is needed to address the role of early posterior effects and expectable subcortical effects in relation to individual levels of fear generalization and particularly its associations with responses to behavioral exposure. An important issue to be addressed by future research is the influence of gender in this interplay. Despite equal distributions of gender-ratios across responders and non-responders, we cannot exclude the possibility that findings might be mainly driven by the higher number of females in our sample. Considering the higher prevalence of animal phobia and anxiety disorders in general in females [40], neurocognitive processes associated with treatment-outcome prediction should be examined with regard to a potential gender specificity.

#### SM2.5. Prediction of individual responses to treatment – an exploratory machine learning approach.

<u>*Results*</u>

Prediction performance marginally exceeded chance level with a mean balanced accuracy of 0.586 (p = 0.076), which varied considerably across the five folds (balanced accuracy SD: 0.139, min: 0.429, max: 0.786). Mean sensitivity and specificity for detecting responder status were 0.643 and 0.529. The mean AUC was 0.58 (see supplemental Figure 1).

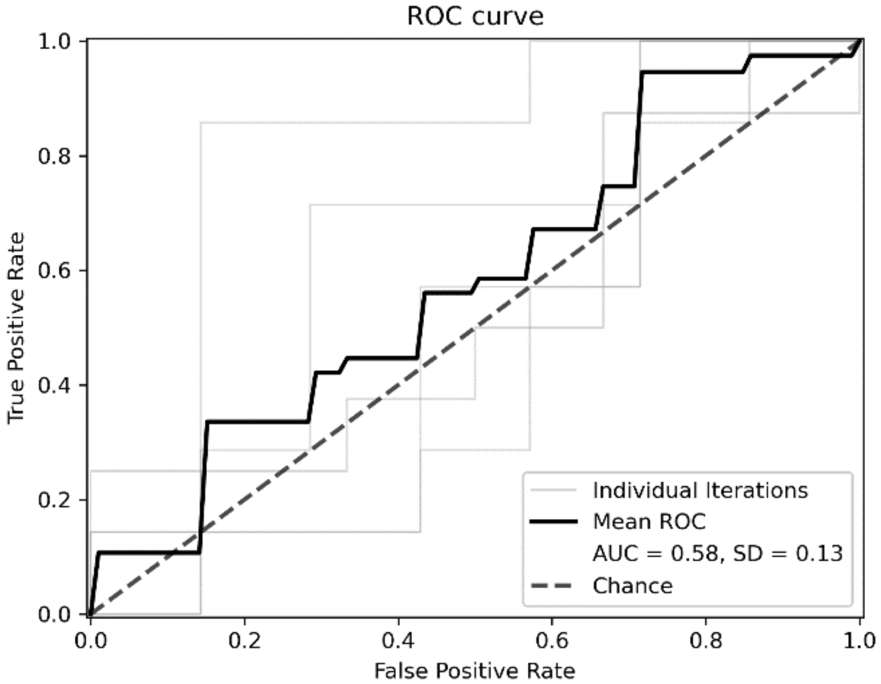


**Figure S7**. Receiver operating characteristic (ROC) curves for the analysis of prediction performance.

#### *Discussion*

We applied machine learning on pre-treatment differences of behavioral and neural markers of fear generalization to predict categorical treatment outcomes (TR-C) on an individual patient level. Given that this analysis was conducted post-hoc, a truly independent split in train and test sets was not possible. We therefore conducted a "conservative" analysis, using all available predictors yielding linear or quadratic generalization gradients, that were identified independently of *treatment-response*. Although, this approach most likely underestimates the true predictive potential of behavioral and neural generalization gradients, the prediction accuracy was comparable to a similar approach on sociodemographic and clinical questionnaire data from in our recent clinical study [23]. Although MEG has already been used to classify subjects according to a clincial diagnosis (e.g. [41], for PTSD), this is the first study to use MEG data for CBT outcome prediction. As expected considering the moderate sample size available in this study, prediction performance varied considerably across the folds of the cross validation procedure. Larger samples for training would be needed to receive more stable predictive models, and larger samples for testing would be needed for a more accurate estimate of prediction performance. The present results demonstrate that behavioral and neural markers of fear generalization may hold valuable information for the prediction of individual CBT outcomes and that further investigation in new and larger samples appears promising.

#### SM3 References

**S**[1] Schwarzmeier H, Leehr EJ, Böhnlein J, et al. Theranostic markers for personalized therapy of spider phobia: Methods of a bicentric external cross-validation machine learning approach. *Int J Methods Psychiatr Res*. Epub ahead of print 8 December 2019. DOI: 10.1002/mpr.1812.

[13] McTeague LM, Gruss LF, Keil A. Aversive learning shapes neuronal orientation

tuning in human visual cortex. *Nat Commun* 2015; 6: 7823.

[23] Leehr EJ, Roesmann K, Böhnlein J, et al. Clinical predictors of treatment response towards exposure therapy in virtuo in spider phobia: a machine learning and external cross-validation approach. *J Anxiety Disord*.
