## supplemental table S1 for "Behavioral and Magnetoencephalographic Correlates of Fear Generalization Are Associated with Responses to Later Virtual Reality Exposure Therapy in Spider Phobia"

**Table S1: Demographic and clinical characteristics of treatment responders and non-responders before and after VRET for the MEG sample**

| Characteristic | Responders [#] | | Non-Responders [##] | | Test | | |
|---|---|---|---|---|---|---|---|
| | M | SD | M | SD | *t [χ²]* | *df* | *p* |
| **Primary Response Criterion** (Reduction in SPQ [%]) | >= 30 | | < 30 | | | | |
| Sample size [N, %] | 36 | 51.42 | 34 | 48.57 | | | |
| Reduction in SPQ [%, SD] | 41.56 | 9.06 | 20.19 | 6.64 | -11.196 | 68 | < .001*** |
| **Demographic Characteristics** | | | | | | | |
| Female gender [N, %] | 29 | 80.56 | 26 | 86.67 | $\chi^2$ = 0.173 | 1 | .677 |
| Age (years) | 26.58 | 8.29 | 30.00 | 8.77 | 1.675 | 68 | .098 |
| Years of education | 14.89 | 2.59 | 14.56 | 3.10 | -0.500 | 67 | .619 |
| **Clinical Characteristics** | | | | | | | |
| Age of onset (years) | 6.26 | 4.72 | 7.09 | 5.64 | 0.665 | 67 | .509 |
| Comorbid major depression [N, %] | 3 | 8.6 | 1 | 2.9 | | | |
| Comorbid subordinate animal phobia [N, %] | 1 | 2.8 | 1 | 2.9 | | | |
| SPQ pre | 22.94 | 2.11 | 22.41 | 2.03 | 1.075 | 68 | .286 |
| SPQ post | 13.36 | 2.14 | 17.88 | 2.18 | 8.745 | 68 | <.001*** |
| BAT (final distance in cm) pre | 172.90 | 79.34 | 171.46 | 65.58 | 0.083 | 68 | .934 |
| BAT (final distance in cm) post | 79.18 | 69.01 | 100.78 | 67.37 | 1.324 | 68 | .190 |
| Reduction in BAT [%, SD] | 55.99 | 32.75 | 40.71 | 30.78 | .1.975 | 66 | .052 |
| CGI pre [N, %] | | | | | $\chi^2$ = 2.480 | 3 | .479 |

| Characteristic | Responders # | | Non-Responders ## | | Test | | |
|---|---|---|---|---|---|---|---|
| | **M** | **SD** | **M** | **SD** | ***t [χ²]*** | ***df*** | ***p*** |
| Mildly ill | 2 | 5.6 | 4 | 11.8 | | | |
| Moderately ill | 21 | 58.3 | 15 | 44.1 | | | |
| Markedly ill | 13 | 36.1 | 12 | 35.3 | | | |
| Severely ill | 0 | 0.0 | 1 | 2.9 | | | |
| CGI post [N, %] | | | | | $\chi^2$ = 8.956 | 4 | .062 |
| Not ill | 2 | 5.6 | 0 | 0.0 | | | |
| Marginally ill | 5 | 13.9 | 1 | 2.9 | | | |
| Mildly ill | 20 | 55.6 | 16 | 47.1 | | | |
| Moderately ill | 6 | 16.7 | 15 | 44.1 | | | |
| Markedly ill | 2 | 5.6 | 2 | 5.9 | | | |
| Severely ill | 0 | 0.0 | 0 | 0.0 | | | |
| STAI trait pre | 35.08 | 10.04 | 36.70 | 8.42 | 0.720 | 67 | .474 |
| BDI-II total pre | 3.90 | 5.05 | 3.85 | 4.39 | 0.035 | 67 | .972 |
| BDI-II total post | 4.17 | 6.09 | 3.21 | 3.51 | 0.802 | 68 | .425 |
| UI-18: total pre | 40.92 | 14.12 | 40.00 | 12.84 | 0.281 | 67 | .779 |
| UI-18: total post | 41.31 | 14.00 | 43.06 | 13.21 | 0.538 | 68 | .592 |

*Note*. Means, standard deviations, t-values (M, SD, t; except where noted), degrees of freedom (df) and significance level (p, two-sided) for each characteristic; SPQ, Spider Phobia Questionnaire (1,2); BAT, behavioral avoidance test; CGI, Clinical Global Impression (3); STAI-Trait, trait-version of the State-Trait Anxiety Inventory (4); BDI-II, Beck Depression Inventory-II (5); UI-18, Intolerance of Uncertainty Scale – 18 (6). # Missing values for responders: Age of onset (1), CGI rating post (1) ## Missing values for non-responders: Years of education (1), CGI rating pre (2), STAI pre (1), BDI pre (1), UI-18: total pre (1); Imputed data for non-responders: BAT pre (1), BAT post (1).
