## supplemental table S2 for "Behavioral and Magnetoencephalographic Correlates of Fear Generalization Are Associated with Responses to Later Virtual Reality Exposure Therapy in Spider Phobia"

**Table S2: Effects of Fear Ratings and UCS-Expectancy Ratings after Test Phase (MEG sample)**

| **Ratings** | **Investigated Type of Treatment Outcome** | **Effect** | **df_n** | **df_d** | ***F*** | ***p*** | $\eta^2$ |
|---|---|---|---|---|---|---|---|
| Fear Rating | Categorical (Anova) | *stimulus-type* (lin) | 1 | 68 | 107.889 | <.001*** | 0.613 |
| | | *stimulus-type* (qu) | 1 | 68 | 3.065 | .085 | 0.043 |
| | | *stimulus-type* * *TR-cat* (lin) | 1 | 68 | 0.156 | .694 | 0.002 |
| | | *stimulus-type* * *TR-cat* (qu) | 1 | 68 | 5.399 | .023* | 0.074 |
| | | *stimulus-type** *TR-cat* * *UCS-type* (lin) | 1 | 68 | 0.420 | .519 | 0.006 |
| | | *stimulus-type** *TR-cat* * *UCS-type* (qu) | 1 | 68 | 0.001 | .976 | 0.000 |
| | | *TR-cat* | 1 | 68 | 0.001 | .976 | 0.000 |
| | | US | 1 | 68 | 31.086 | .000 | 0.314 |
| | | *UCS-type** *TR-cat* | 1 | 68 | 0.759 | .387 | 0.011 |
| | | *UCS-type** *stimulus-type* (lin) | 1 | 68 | 11.199 | .001*** | 0.141 |
| | | *UCS-type** *stimulus-type* (qu) | 1 | 68 | 1.223 | .273 | 0.018 |
| | Dimensional (Ancova) | *stimulus-type* * *TR-dim* (lin) | 1 | 68 | 0.048 | .828 | 0.001 |
| | | *stimulus-type** *TR-dim* (qu) | 1 | 68 | 3.099 | .083[t] | 0.044 |
| | | *stimulus-type** *TR-dim* * *UCS-type* (lin) | 1 | 68 | 0.116 | .735 | 0.002 |
| | | *stimulus-type** *TR-dim* * *UCS-type* (qu) | 1 | 68 | 0.023 | .880 | 0.000 |
| | | *UCS-type** *TR-dim* | 1 | 68 | 1,016 | .317 | 0.015 |
| Expectancy ratings | Categorical (Anova) | *stimulus-type* (lin) | 1 | 68 | 207.777 | <.001*** | 0.753 |
| | | *stimulus-type* (qu) | 1 | 68 | 28.074 | <.001*** | 0.292 |
| | | *stimulus-type* * *TR-cat* (lin) | 1 | 68 | 0.867 | .355 | 0.013 |

| **Ratings** | **Investigated Type of Treatment Outcome** | **Effect** | **df_n** | **df_d** | ***F*** | ***p*** | $\eta^2$ |
|---|---|---|---|---|---|---|---|
| | | *stimulus-type* * *TR-cat* (qu) | 1 | 68 | 0.305 | .582 | 0.004 |
| | | *stimulus-type** *TR-cat* * *UCS-type* (lin) | 1 | 68 | 0.011 | .917 | 0.000 |
| | | *stimulus-type** *TR-cat* * *UCS-type* (qu) | 1 | 68 | 0.540 | .465 | 0.008 |
| | | *TR-cat* | 1 | 68 | 0.127 | .722 | 0.002 |
| | | US | 1 | 68 | 2.320 | .132 | 0.033 |
| | | *UCS-type** *TR-cat* | 1 | 68 | 2.163 | .146 | 0.031 |
| | | *UCS-type** *stimulus-type* (lin) | 1 | 68 | 1.276 | .263 | 0.018 |
| | | *UCS-type** *stimulus-type* (qu) | 1 | 68 | 0.018 | .895 | 0.000 |
| | Dimensional (Ancova) | *stimulus-type* * *TR-dim* (lin) | 1 | 68 | 0.177 | .676 | 0.003 |
| | | *stimulus-type** *TR-dim* (qu) | 1 | 68 | 0.785 | .379 | 0.011 |
| | | *stimulus-type** *TR-dim* * *UCS-type*(lin) | 1 | 68 | 0.006 | .940 | 0.000 |
| | | *stimulus-type** *TR-dim* * *UCS-type*(qu) | 1 | 68 | 0.608 | .438 | 0.009 |
| | | *UCS-type** *TR-dim* | 1 | 68 | 1.142 | .289 | 0.017 |
