## supplemental table S3 for "Behavioral and Magnetoencephalographic Correlates of Fear Generalization Are Associated with Responses to Later Virtual Reality Exposure Therapy in Spider Phobia"

**Table S3: Effects within clusters revealing significant linear main effects. We tested for modulatory effects of treatment outcomes (*TR-cat*, *TR-dim*) on linear and quadratic gradients**

| Analysis | Cluster | Investigated Type of Treatment Outcome | Effect | df_n | df_d | F | p | η2 |
|---|---|---|---|---|---|---|---|---|
| Anterior, 0-300ms | Ant-Neg-1 (110-157ms)* | Categorical (Anova) | *stimulus-type* * *TR-cat* (lin) | 1 | 68 | 9.351 | .003** | 0.121 |
| | | | *stimulus-type* * *TR-cat* (qu) | 1 | 68 | 0.155 | .695 | 0.002 |
| | | Dimensional (Ancova) | *stimulus-type* * *TR-dim* (lin) | 1 | 68 | 6.616 | .012* | 0.089 |
| | | | *stimulus-type* * *TR-dim* (qu) | 1 | 68 | 0.125 | .724 | 0.002 |
| | Ant-Neg-2 (210-300ms)* | Categorical (Anova) | *stimulus-type* * *TR-cat* (lin) | 1 | 68 | 1.485 | .227 | 0.021 |
| | | | *stimulus-type* * *TR-cat* (qu) | 1 | 68 | 1.006 | .319 | 0.015 |
| | | Dimensional (Ancova) | *stimulus-type* * *TR-dim* (lin) | 1 | 68 | 1.321 | .254 | 0.019 |
| | | | *stimulus-type* * *TR-dim* (qu) | 1 | 68 | 7.499 | .008** | 0.099 |
| | Ant-Neg-3 (153-230ms) | Categorical (Anova) | *stimulus-type* * *TR-cat* (lin) | 1 | 68 | 0.410 | .524 | 0.006 |
| | | | *stimulus-type* * *TR-cat* (qu) | 1 | 68 | 0.158 | .692 | 0.002 |
| | | Dimensional (Ancova) | *stimulus-type* * *TR-dim* (lin) | 1 | 68 | 1.149 | .288 | 0.017 |
| | | | *stimulus-type* * *TR-dim* (qu) | 1 | 68 | 0.059 | .809 | 0.001 |
| | Ant-Neg-4 (180-240ms) | Categorical (Anova) | *stimulus-type* * *TR-cat* (lin) | 1 | 68 | 1.271 | .263 | 0.018 |
| | | | *stimulus-type* * *TR-cat* (qu) | 1 | 68 | 0.005 | .944 | 0.000 |
| | | Dimensional (Ancova) | *stimulus-type* * *TR-dim* (lin) | 1 | 68 | 2.033 | .158 | 0.029 |
| | | | *stimulus-type* * *TR-dim* (qu) | 1 | 68 | 0.046 | .831 | 0.001 |
| Anterior, 300-600ms | - | - | - | - | - | - | - | - |
| Posterior, 300-600ms | Post-Pos-1 (437-567ms) | Categorical (Anova) | *stimulus-type* * *TR-cat* (lin) | 1 | 68 | 0.412 | .523 | 0.006 |

| **Analysis** | **Cluster** | **Investigated Type of Treatment Outcome** | **Effect** | **df_n** | **df_d** | **F** | **p** | **η2** |
|---|---|---|---|---|---|---|---|---|
| | | | *stimulus-type* * *TR-cat* (qu) | 1 | 68 | 1.345 | .250 | 0.019 |
| | | Dimensional (Ancova) | *stimulus-type* * *TR-dim* (lin) | 1 | 68 | 0.795 | .376 | 0.012 |
| | | | *stimulus-type* * *TR-dim* (qu) | 1 | 68 | 0.631 | .430 | 0.009 |
| Posterior, 0-300 ms | Post-Neg-1 (47-100ms) | Categorical (Anova) | *stimulus-type* * *TR-cat* (lin) | 1 | 68 | 4.057 | .048* | 0.056 |
| | | | *stimulus-type* * *TR-cat* (qu) | 1 | 68 | 0.078 | .780 | 0.001 |
| | | Dimensional (Ancova) | *stimulus-type* * *TR-dim* (lin) | 1 | 68 | 4.003 | .049* | 0.056 |
| | | | *stimulus-type* * *TR-dim* (qu) | 1 | 68 | 0.096 | .758 | 0.001 |

* clusters presented in the main text
