## supplemental table S4 for "Behavioral and Magnetoencephalographic Correlates of Fear Generalization Are Associated with Responses to Later Virtual Reality Exposure Therapy in Spider Phobia"

### Table S4. Effects in clusters resulting from orthogonal contrasts.

| Analysis | Cluster | Investigated Type of Treatment Treatment Outcome | Effect | df_n | df_d | *F* | *P* | $\eta^2$ |
|---|---|---|---|---|---|---|---|---|
| Anterior, 0-300ms | - | - | - | - | - | - | - | - |
| Anterior, 300-600ms | Ant-Int-1 (507-557ms)* | Categorical (Anova) | *stimulus-type* (lin) | 1 | 68 | 0.301 | .184 | 0.004 |
| | | | *stimulus-type* (qu) | 1 | 68 | 0. 241 | .625 | 0.004 |
| | | | *stimulus-type* * *TR-cat* (lin) | 1 | 68 | 12.829 | .001** | 0.159 |
| | | | NR: *stimulus-type* (lin) | 1 | 33 | 10.698 | .003** | 0.245 |
| | | | R: *stimulus-type* (lin) | 1 | 35 | 3.907 | .056 | 0.100 |
| | | | *stimulus-type* * *TR-cat* (qu) | 1 | 68 | 0.352 | .555 | 0.005 |
| | | | *stimulus-type** *TR-cat* * US (lin) | 1 | 68 | 0.211 | .648 | 0.003 |
| | | | *stimulus-type** *TR-cat* * US (qu) | 1 | 68 | 2.267 | .137 | 0.032 |
| | | | US | 1 | 68 | 1.803 | .184 | 0.026 |
| | | | *UCS-type* * *TR-cat* | 1 | 68 | 0.438 | .510 | 0.006 |
| | | | *UCS-type* * *stimulus-type* (lin) | 1 | 68 | 0.007 | .934 | 0.000 |
| | | | *UCS-type* * *stimulus-type* (qu) | 1 | 68 | 0.086 | .770 | 0.001 |
| | | Dimensional (Ancova) | *stimulus-type* * *TR-dim* (lin) | 1 | 68 | 6.498 | .013* | 0.087 |
| | Ant-Int-2 (427-493 ms)* | Categorical (Anova) | *stimulus-type* (lin) | 1 | 68 | 0.258 | .552 | 0.005 |
| | | | *stimulus-type* (qu) | 1 | 68 | 0.068 | .795 | 0.001 |
| | | | *stimulus-type* * TR (lin) | 1 | 68 | 19.734 | <.001*** | 0.225 |
| | | | NR: *stimulus-type* (lin) | 1 | 33 | 7.993 | .008** | 0.195 |
| | | | R: *stimulus-type* (lin) | 1 | 35 | 11.937 | .001** | 0.254 |

| **Analysis** | **Cluster** | **Investigated Type of Treatment Treatment Outcome** | **Effect** | **df_n** | **df_d** | ***F*** | ***P*** | ***η²*** |
|---|---|---|---|---|---|---|---|---|
| | | | *stimulus-type* * TR (qu) | 1 | 68 | 0.000 | .987 | 0.000 |
| | | | *stimulus-type** TR * US (lin) | 1 | 68 | 0.002 | .967 | 0.000 |
| | | | *stimulus-type** TR * US (qu) | 1 | 68 | 0.096 | .758 | 0.001 |
| | | | US | 1 | 68 | 1.992 | .163 | 0.028 |
| | | | *UCS-type* * TR | 1 | 68 | 0.061 | .806 | 0.001 |
| | | | *UCS-type* * *stimulus-type* (lin) | 1 | 68 | 0.750 | .389 | 0.011 |
| | | | *UCS-type* * *stimulus-type* (qu) | 1 | 68 | 0.713 | .401 | 0.010 |
| | | Dimensional (Ancova) | *stimulus-type* * *TR-dim* (lin) | 1 | 68 | 15.901 | <.001*** | 0.190 |
| Posterior, 0-300ms | Post-Int-1 (130-160 ms) | Categorical (Anova) | *stimulus-type* (lin) | 1 | 68 | 0.064 | .801 | 0.001 |
| | | | *stimulus-type* (qu) | 1 | 68 | 0.037 | .848 | 0.001 |
| | | | *stimulus-type* * TR (lin) | 1 | 68 | 13.462 | <.001*** | 0.165 |
| | | | NR: *stimulus-type* (lin) | 1 | 33 | 7.435 | .010* | 0.184 |
| | | | R: *stimulus-type* (lin) | 1 | 35 | 6.472 | .016* | 0.156 |
| | | | *stimulus-type* * TR (qu) | 1 | 68 | 3.223 | .077 | 0.045 |
| | | | *stimulus-type** TR * US (lin) | 1 | 68 | 0.003 | .956 | 0.000 |
| | | | *stimulus-type** TR * US (qu) | 1 | 68 | 0.006 | .938 | 0.000 |
| | | | US | 1 | 68 | 0.022 | .881 | 0.000 |
| | | | *UCS-type* * TR | 1 | 68 | 0.503 | .481 | 0.007 |
| | | | *UCS-type* * *stimulus-type* (lin) | 1 | 68 | 0.009 | .926 | 0.000 |
| | | | *UCS-type* * *stimulus-type* (qu) | 1 | 68 | 0.075 | .785 | 0.001 |

| **Analysis** | **Cluster** | **Investigated Type of Treatment Treatment Outcome** | **Effect** | **df_n** | **df_d** | ***F*** | ***P*** | ***$\eta^2$*** |
|---|---|---|---|---|---|---|---|---|
| | | Dimensional (Ancova) | *stimulus-type* * *TR-dim* (lin) | 1 | 68 | 4.519 | .037* | 0.062 |
| Posterior, 300-600ms | - | | - | - | - | - | - | - |

* clusters presented in the main text
